# Assessing the Impact of Non-Pharmaceutical Interventions on COVID-19: A Combined CCE and Quantile Regression Approach

**DOI:** 10.1101/2023.11.27.23299097

**Authors:** Kaibalyapati Mishra

## Abstract

This paper tries to quantify the impact of government policy intervention on the death due to COVID-19 in India. I use the Oxford COVID-19 Government Response Tracker (OxCGRT), a longitudinal database of daily government response from Jan 28th, 2020, when the first COVID case was diagnosed in India till December 31st, 2022. Here government responses are captured in form of, *stringency* measures, *containment* measures, *economic support* measures, and the *overall government support*, providing a holistic assessment of the government’s efforts in mitigating the virus’s incidence. I quantify both the average relation and causality at the to understand the impacts of NPIs with COVID-19 incidence in terms of deaths and infections due to COVID-19. Short-term analysis reveals a significant relationship between various non-pharmaceutical interventions (NPIs) and the logarithmic change in COVID-19 deaths. Higher infection rates are strongly associated with increased deaths, with positive coefficients. Stringent measures, containment actions, and economic support show negative coefficients, indicating that these interventions effectively reduce deaths in the short term. The overall government support, which aggregates all three NPIs, also demonstrates a significant negative effect on deaths, highlighting the importance of stringent and immediate measures in controlling the death toll early in the pandemic. In the long term, the analysis continues to emphasize the importance of infection rates and NPIs. Long-term coefficients for infection rates and various NPIs are consistently significant and negative, indicating that sustained interventions significantly reduce mortality over time. Specific measures like stringency, containment, and economic support show substantial negative impacts underscoring the long-term benefits of maintaining rigorous public health measures. Further, causality analysis confirms that relationship among government interventions and COVID-19 incidences were mostly bidirectional, meaning more deaths (or infections) leads to stricter interventions that in turn further reduce deaths.

**JEL:** *C23, C54, I18, I38*

## 1 Introduction

The COVID-19 pandemic posed a significant challenge to public health authorities worldwide, prompting governments to implement various non-pharmaceutical interventions (NPIs) to mitigate the spread of the virus. Government responses to COVID-19 represent some of the most globally impactful events of the 21st century. The extent to which these responses, including measures like school closures, were associated with changes in COVID-19 outcomes remains a topic of ongoing debate. The emergence of the COVID-19 pandemic raised a multitude of questions that demanded contributions from various fields of science and policy-making. Scientific and health experts played a crucial role in developing vaccines and implementing infection control measures. However, the responsibility for vaccine distribution, the implementation of restrictions, and the preservation of livelihoods ultimately fell on one entity: *the government*.

In a nation like India, poised as one of the world’s fastest-growing economies^1^ and on the cusp of surpassing China in terms of population, addressing the havoc wrought by the pandemic demanded systematic and well-coordinated actions from both state and central governments. The population prediction or India and specific composition of its demographic groups impacts its route for mitigating the COVID-19 challenge distinctively. As of January 27, 2023, India reported over 45 million COVID cases, with 0.1% of these cases resulting in fatalities (over 0.5 million deaths), making India second only to the USA in total deaths. However, India’s initial testing efforts paint a different picture. Despite ranking second in total tests conducted globally, India was only 12th in tests conducted per million people during the same time. Furthermore, India’s vast geographical and socio-economic diversity compelled the imposition of both nationwide and region-specific policies. In this context, this paper seeks to analyze the government’s response to COVID-19 in India and its various states, with a focus on their targeted policy responses and an economic evaluation.

The spread of COVID-19 was characterized by its high infectivity, especially in countries like India with dense populations (Shereen, Khan, Kazmi, Bashir, & Siddique, 2020). The virus’s higher infectiousness, coupled with increased mortality rates across its variants (Wang et al., 2020), had a profound impact on the economic, social, and psychological aspects of life. The graph in Figure-1 vividly portrays the persistence of infection, the evolution of new case trends (waves), and their eventual decline over time. This graph also highlights the positive effect of collective efforts such as vaccination, social distancing, and lock-downs by the government on reducing peak infection spikes and shortening the duration of waves. While almost similar trend is followed by deaths, it plausible to claim that deaths have declined gradually post September 2021. Nevertheless, India’s testing efforts have been relatively modest in comparison to its vast population (Thiagarajan, 2021).

### 1.1 COVID-19: The Indian COVID-19 Chronicle

The first of the novel coronavirus (COVID-19) was recorded in Wuhan, China, in December 2019. Swift transmission of the virus ensued, affecting a significant number of people within a month. India’s first reported case was reported in late January 2020 in the state of Kerala, with the individual having recently returned from China. Following this, there has been a notable upswing in COVID-19 cases across various Indian states. In recognition of the gravity of the situation, the government initiated a 21-day nationwide lockdown from March 25, 2020, to April 14, 2020. This stringent measure aimed to mitigate the spread of COVID-19, resulting in the closure of industries, academic institutions, markets, and public gatherings. Subsequent to the initial lockdown, three successive lockdown phases were implemented (April 15 to May 3, 2020; May 4 to May 17, 2020; May 18 to May 31, 2020). In an effort to revive the Indian economy, two unlock phases were subsequently introduced (June 1 to June 30, 2020, and July 1 to July 31, 2020), as shown in figure-1.

A detailed depiction of the government response in India has been pictorially presented in Figure-6a. On March 29, 2020, the Government of India established 11 empowered groups to address various aspects of COVID-19 management in the country. These groups were tasked with making informed decisions on a wide range of issues, including medical emergency planning, hospital availability, isolation and quarantine facilities, disease surveillance and testing, essential medical equipment availability, human resource and capacity building, supply chain and logistics management, coordination with the private sector, economic and welfare measures, information, communications, and public awareness, technology and data management, public grievances, and strategic issues related to lockdown. On September 10, 2020, these groups underwent restructuring to adapt to the changing needs and evolving scenario.

The Ministry of Health & Family Welfare (MoFHW) unveiled containment strategies to address both cluster and widespread outbreaks on March 2nd and April 4th, 2020, respectively. These plans underwent periodic updates. The containment strategies focus on disrupting the transmission chain through *(i) delineating containment and buffer zones, (ii) implementing stringent perimeter control, (iii) conducting thorough house-to-house searches for cases and contacts, (iv) isolating and testing suspected cases and high-risk contacts, (v) quarantining high-risk contacts, (vi) intensifying risk communication to enhance community awareness regarding simple preventive measures and the importance of prompt treatment seeking, and (vii) reinforcing passive surveillance for Influenza Like Illness (ILI) and Severe Acute Respiratory Illness (SARI) in containment and buffer zones*.

A system of health facilities consisting of *three tiers* has been established to effectively handle COVID-19 cases. This included: (i) COVID Care Centers equipped with isolation beds for mild or pre-symptomatic cases; (ii) Dedicated COVID Health Centers (DCHCs) providing oxygen-supported isolation beds for moderate cases; and (iii) Dedicated COVID Hospitals (DCHs) with ICU beds for severe cases. Additionally, tertiary care hospitals affiliated with organizations such as ESIC, Defence, Railways, paramilitary forces, and the Steel Ministry have been utilized for the management of cases. Further, instructions for the clinical management of COVID-19 were released, consistently revised, and widely disseminated. These guidelines encompassed various aspects, such as defining cases, implementing infection control measures, conducting laboratory diagnoses, initiating early supportive therapy, addressing severe cases, and managing complications. Furthermore, allowances for investigation therapies, including Remdesivir, Convalescent plasma, and Tocilizumab, were outlined for the treatment of severe cases under rigorous medical supervision. It is important to note that the indices of government response, used as major policy intervention variables, represent a combination of measures for quantification.

### 1.2 Related Literature

The literature studying the impact of the degree of government response is scarce both in general and also in the case of India. In their study Mukherjee, Banerjee, Mitra, and Mukherjee (2022), the desired needs of the hour during the pandemic. They supported the government’s stringent measures in response to COVID-19, such as work-from-home and stringent lock-downs, while also suggesting the need for timely interventions. As attributed earlier, India presents a compelling landscape for the study of non-pharmaceutical interventions (NPIs) in the context of COVID-19 due to its diverse population and geographical variations. With a population exceeding 1.3 billion people, the country’s demographic mosaic offers a unique opportunity to examine the differential impact of interventions across various cultural and regional settings. High population density, particularly in urban areas (Nijman, 2012), contributes to the rapid transmission of infectious diseases, making it imperative to understand how NPIs operate in densely populated regions. Moreover, India exhibits significant variability in healthcare infrastructure (Chaturvedi et al., 2023), economic conditions, and socio-demographic factors, influencing the implementation and effectiveness of NPIs.

The nation has implemented a range of interventions, including lock-downs and social distancing measures, providing a rich data-set (OXCGRT in this case) for analyzing policy implementation and compliance. Given the prevalence of infectious diseases and varying co-morbidities, examining the interplay between pre-existing health conditions and the outcomes of COVID-19 interventions in India contributes valuable insights to the global understanding of pandemic management. India’s global significance further underscores its importance as a lucrative study area for researchers exploring the impact of NPIs on COVID-19.

Herby, Jonung, and Hanke (2023) show several key studies to evaluate the impact of lockdowns on COVID-19 mortality. Björk, Mattisson, and Ahlbom (2021) examines the impact of winter holidays and government responses on mortality across Europe during the first wave of the pandemic, highlighting the varying effects of different policy measures on mortality rates. Similarly, (Bjørnskov, 2021) provides a cross-country comparison of the effectiveness of lockdowns, offering an economic perspective on the lockdown measures’ outcomes. Blanco, Emrullahu, and Soto (2020) contribute to the discussion presenting worldwide evidence on the effectiveness of coronavirus containment measures, which underscores the global variations in policy impacts. Bollyky et al. (2022) analyze pandemic preparedness and its correlation with infection and fatality rates, providing a broad context of how different countries’ preparedness levels influenced their outcomes during the pandemic. (Bonardi, Gallea, Kalanoski, & Lalive, 2020) discuss the local and immediate impacts of lockdown policies on the spread and severity of COVID-19, while Bongaerts, Mazzola, and Wagner (2021) explore the mortality impact of business closures during the pandemic, emphasizing the economic trade-offs of lockdown measures. Brauner et al. (2021) offer insights into the effectiveness of various government interventions against COVID-19, employing robust methodologies to infer causality. Chernozhukov, Kasahara, and Schrimpf (2021) focus on the causal impact of masks, policies, and behavior during the early stages of the pandemic in the U.S., adding a nuanced understanding of different mitigation strategies.

In the context of assessing the impact of NPIs in India I use the confirmed number of deaths as a dependent variable for my analysis. Similarly, I use the degree of government response in terms of stringency measures, containment measures, and overall government support to reduce the infection rate in India as the independent variables. This constitutes a strongly balanced panel but with gaps, that uses data from all the states (28) and union territories (5)^2^ of India. I use daily data on the mentioned dependent and independent variables over almost two years, spanning from 28th January 2021^3^ until 31st December 2022. I use the sub-national (OxCGRT) database on the degree of government responses for all the indices (independent variable) and daily confirmed deaths and infections (dependent variable) from the Ritchie, Roser, and Rosado (2020) database.

The findings from this study suggest robust evidence of effectiveness of NPIs in controlling and curbing deaths in both short and long-term. This is found to be individually true for all the NPIs i.e. stringency, containment and economic support and overall government response that encapsulates the earlier three. I find that, while NPIs are ineffective in controlling infection in the short-term, in the long they are promptly successful. Further we find that quantile regression not only confirms the robustness of the earlier presented findings, it also unveils the heterogeneity across quantiles. It confirms that NPIs are initially ineffective in controlling infection however over the longer-term they have successfully cubed it. From, causality analysis we confirm that, the relationship between NPIs and incidences of COVID-19 are both bi-directional and significant.

The subsequent sections are organized as follows: In §2, I delve into the government response metrics utilized for the analysis and methodology aspect of the paper. §3 presents the results and discusses the findings of the study. §4 outlines the major policy suggestions, while §5 concludes.

## 2 Data and Methodology

For this paper, I use data from two sources. *First,* I use the India: Coronavirus Pandemic Country Profile data (Ritchie, Roser, & Rosado, 2022) for the weekly data on COVID-19-related deaths and infections, which are the dependent variables. *Second,* I also use the Oxford COVID-19 Government Response Tracker (OxCGRT) database (Hale et al., 2021) for the independent variables that encompass the non-pharmaceutical interventions (NPIs) by the Government of India (GoI) since the inception of the COVID-19 pandemic in India. Table-5 summarizes the key descriptive statistics of these variables. To address the skewness in the raw data, I have applied a logarithmic transformation to all the variables.The table 6 represents the matrix of correlation among variables. From here, it can be observed that the independent variables, i.e., LnSTRINGENCY, LnCONTAINMENT, LnECONOMIC, and LnOVERALL, are highly correlated. This is because the steps taken by the government are supply-side measures, and the analysis does not involve any demand-side measures or actions taken by the residents on their own. Thus, the impact of each of these measures on the number of deaths and infections cannot be claimed to be due to exclusive measures, as all three measures (which are combined in LnOVERALL) were active during each of the waves. Assuming that there will be overlapping impacts, we use them separately in different equations to quantify the average association of respective NPI. I discuss further on the variables in section-2.1.

### 2.1 Variables and framework

The figure-2 outlines the detailed framework consisting of the dependent and independent variables of the study, objectives, methods, and the scheme of estimation procedures used in this paper. The COVID-19 pandemic necessitated a variety of interventions from the government. Along with pharmaceutical interventions like vaccination, several non-pharmaceutical interventions (NPIs) were implemented. Moreover, since the preparation, production, and disbursement of vaccines took a considerable amount of time initially, NPIs were the significant government interventions that directly and indirectly impacted deaths and infections due to COVID-19 in India. The NPIs included both targeted and general interventions, which at times varied across geographic and sectoral dimensions. The OxCGRT database is a novel endeavor in this regard because it captures these interventions and forms indices under the categories of stringency index (LnSTRINGENCY), containment index (LnCONTAINMENT), economic support index (LnECONOMIC), and overall government support index (LnOVERALL). LnOVERALL encompasses all the interventions. We use them in separate equations to identify the respective average association of each of the indicators. Table-4 provides detailed sub-indices of each of the NPIs used in this paper.

Hale et al. (2020) outline that governments’ responses to COVID-19 show significant variation. For example, C1^4^ (school closing) varies widely: some places shut all schools, others close universities and primary schools at different times, and some keep schools open for children of essential workers. These differences, influenced by local political and social contexts, make systematic comparison challenging. Composite measures, while simplifying data, may overlook nuances. They help reduce complexity and prevent over-interpretation of individual indicators but can miss critical details and make assumptions about relevance. Each index (independent variable here) consists of individual policy response indicators. OxCGRT creates a score for each indicator by subtracting half a point if the policy is general instead of targeted, then rescales it to a 0-100 range. These scores are averaged to form the composite indices. The figure 3 demonstrates the variation of the daily index values in India. As visible, index values of NPIs don’t vary on a daily basis; rather, they remain the same for some time. Due to this, in our independent variables (as shown in figure 2), we take weekly averages of the NPIs.

**Figure 1:**
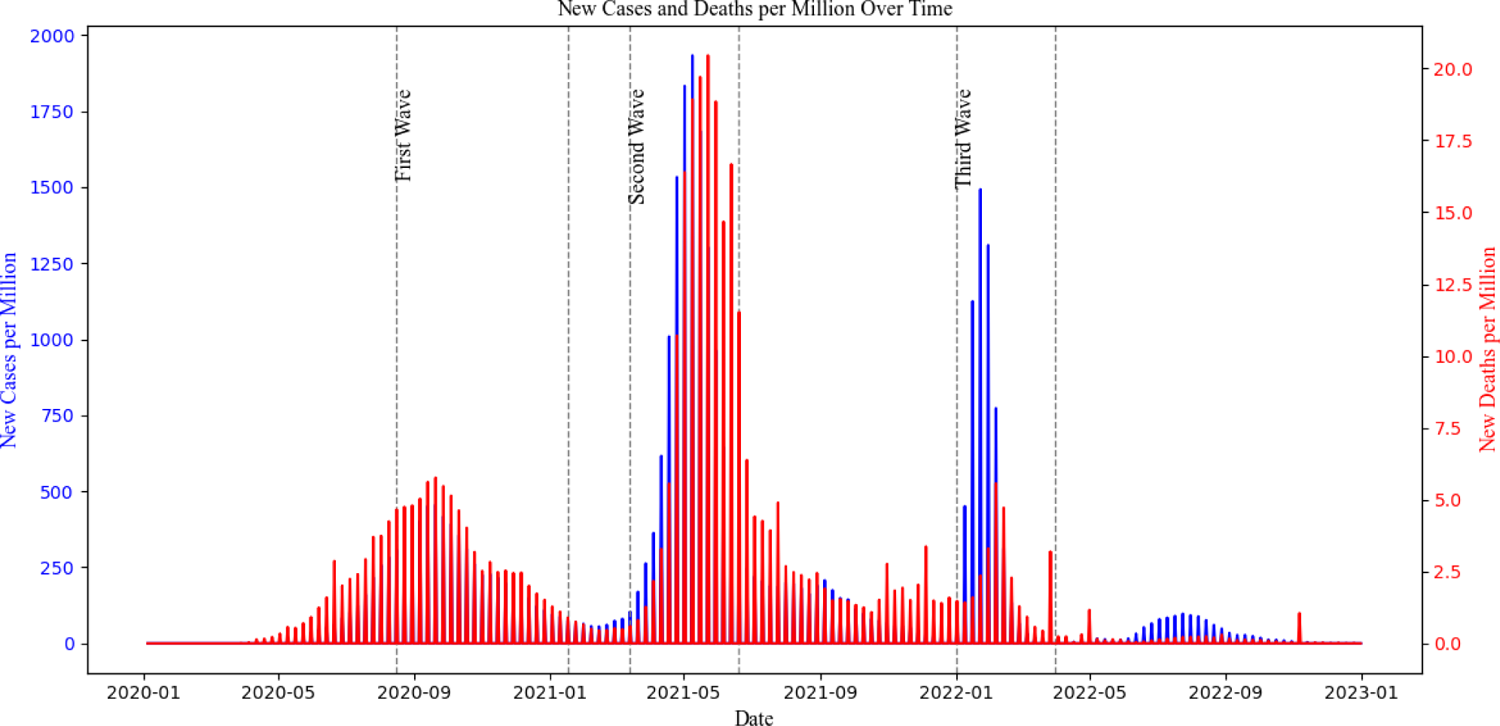
New cases and deaths per million population over time from 2020 to 2023

**Figure 2:**
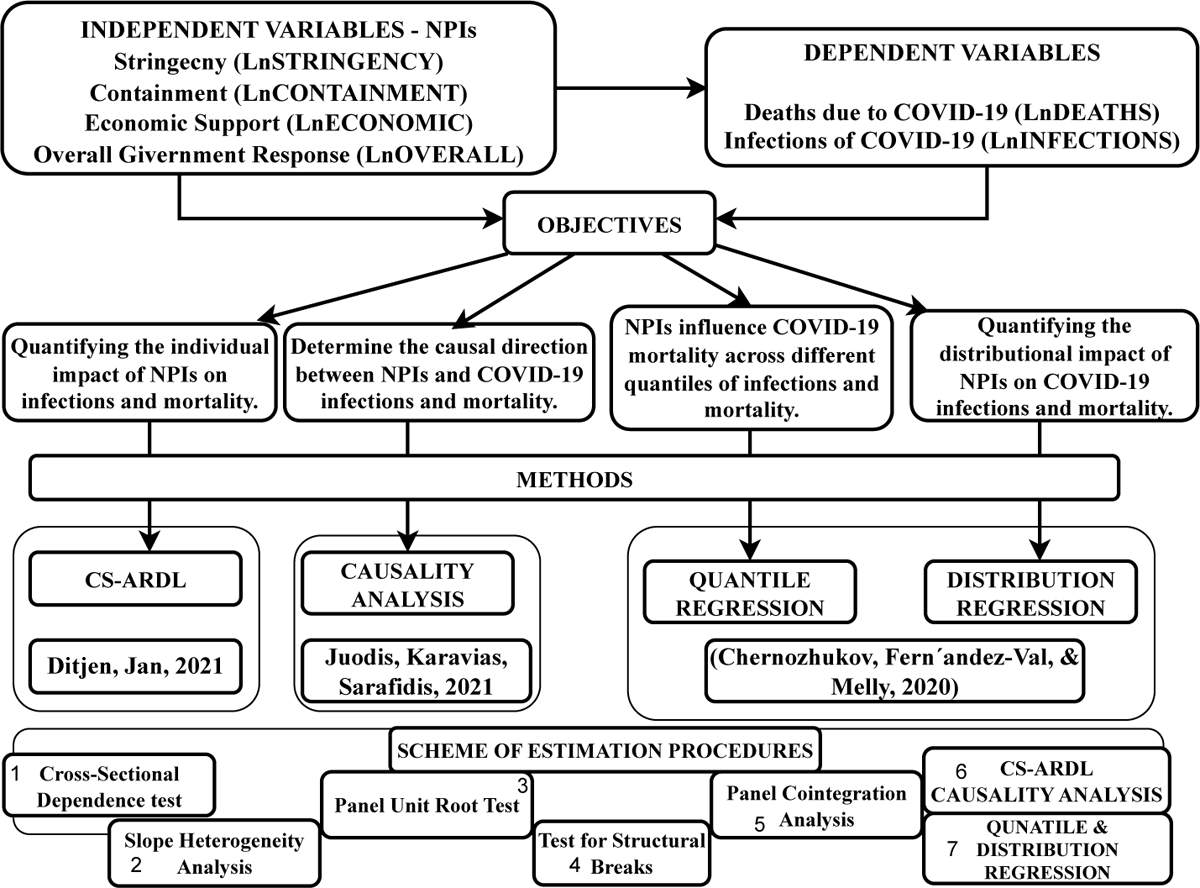
The detailed framework of the study outlining the dependent and independent variables, objectives, methods, and the scheme of estimation procedures used.

**Figure 3:**
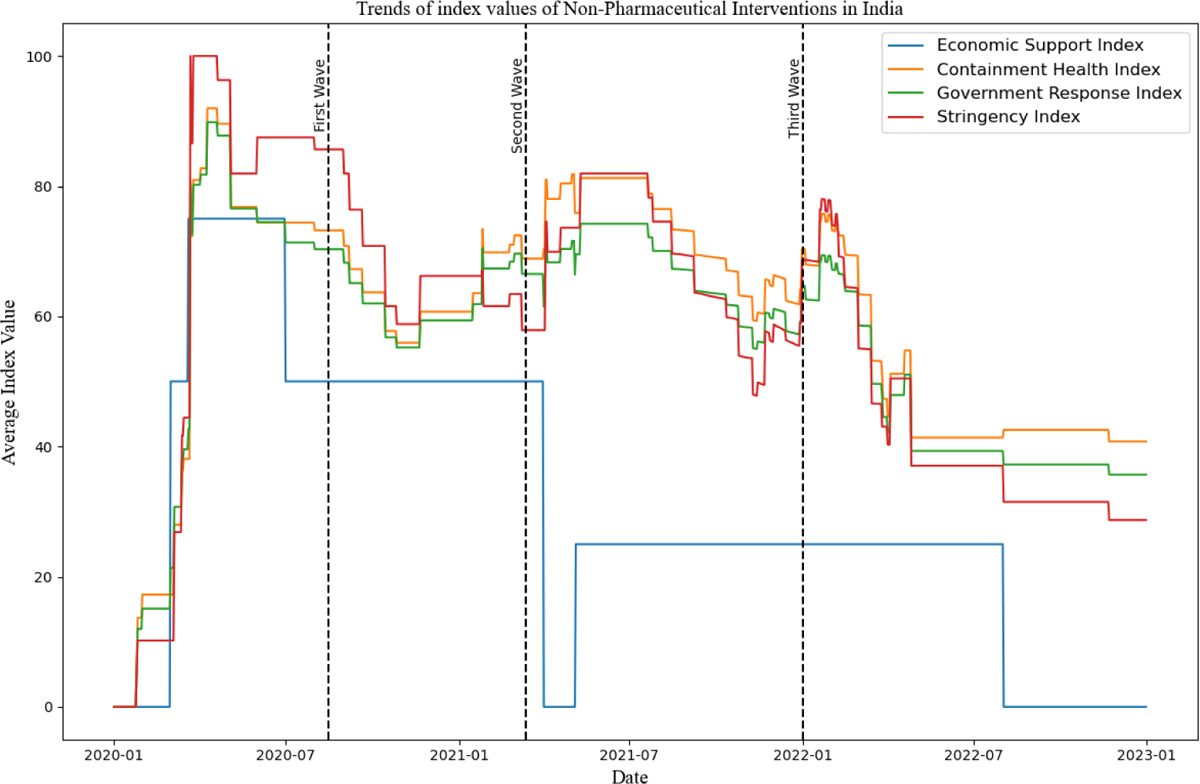
Line graphs demonstrating the trends of all NPIs from 2020 to 2022.

*Stringency and containment measures (STI)*, as given in table-4, along with *Containment measures (CNI)* and *Overall government support measures (GRI)*, have common sub-indices of school, public transport, and workplace closure, cancellation of public events, public gathering restrictions, stay-at-home requirements, and internal and international travel restrictions. Along with these, CNI (LnCONTAINMENT) includes international support, information campaigns, health systems, emergency investment in healthcare and in COVID-19 vaccines, and face coverings. However, STI (LnSTRINGENCY) includes only international support over the common indicators mentioned earlier. Meanwhile, GRI (LnOVERALL) includes all the sub-indices that constitute CNI, STI, and ESI (LnECONOMIC). *Economic Support Measures* (ESI) comprise only income support and debt relief.

### 2.2 Econometric Model

This paper examines the average relationship between COVID-19-related deaths and infections with non-pharmaceutical interventions (NPIs) in India. To assess the impact of NPIs efficiently, separate identification equations are provided and estimated accordingly. Additionally, we explore how this relationship varies across quantiles and distributions, ensuring robustness across different analytical perspectives. Thus, I quantify both the average relation and causality at the aggregated level of variables, and also examine the relationship across disaggregated quantiles. These analyses not only provide insights into the average and disaggregated associations of NPIs with COVID-19 incidence, but also enhance the robustness and reliability of our findings. For the state *i* at time *t*, Incidence of COVID-19 pandemic LnDEATHS can be expressed as the function below:

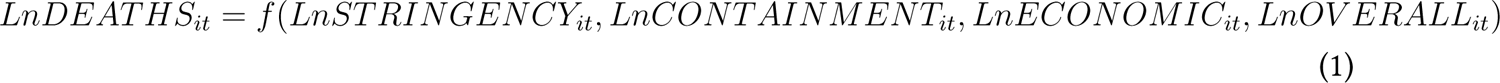

Where i = 1, 2,… N indicates the sampled states, t = 1, 2,. T weeks. Variable names and definitions are available in table-5. Eq. (1) can be rewritten as follows using the logarithmic expression of the variables:

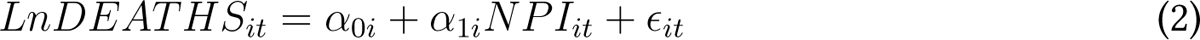

For *α*_0*i*_ is the static impact, *α*_1*i*_ is the coefficients to be estimated, while *ɛ_it_* is the residual. Similar expressions of eq-(1) and (2) are be used with *LnINFECTIONS_it_* being the dependent variables. Following the patter in Namahoro, Wu, and Su (2023), in the coming sub-sections we discuss about a series of pre-testing that has been conducted as given in the scheme of estimation procedures in figure-2, along with their identification as a pretext to discussion in results.

#### 2.2.1 Cross section dependency test

Pesaran (2003) developed Lagrange Multiplier (LM) and CD4 cross-sectional dependence tests tailored for large panel datasets, with subsequent refinements by Pesaran (2015) and Pesaran (2021). In contrast, Breusch and Pagan (1980) introduced the Breusch-Pagan LM test to assess cross-sectional dependence in smaller panel datasets. The exponent of cross-sectional dependence, proposed by Bailey, Kapetanios, and Pesaran (2016), quantifies the strength of relationships identified by the Pesaran and Breusch tests. A simple consistency estimate of this exponent is given by:

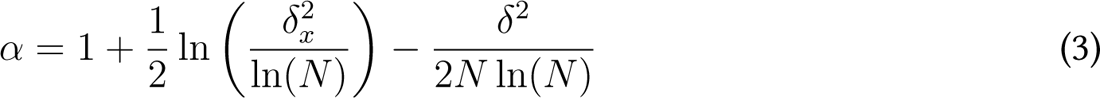

where *α* denotes the alpha exponent of cross-sectional dependence, *δ*^2^ is the variance of the tested variable, and *N*represents the variable size. The value of *α* ranges between 0 and 1, reflecting the strength of relationships. Based on these thresholds, Chudik, Pesaran, and Tosetti (2011) categorizes cross-sectional dependence into four levels: weak (*α* = 0), semi-weak (0 *< α <* 0.5), semi-strong (0.5 *≤ α <* 1), and strong (*α* = 1). The results of the tests are given in table-7.

#### 2.2.2 Panel test for slope heterogeneity

Building on a standardized version of Swamy (1970)’s test, Pesaran and Yamagata (2008) introduced a test for slope homogeneity suitable for panel data with large N and T. The standard delta test for slope heterogeneity in large panels as suggested by Pesaran and Yamagata (2008) involves the following statistics. The Δ statistic and the adjusted Δ statistic, which adjusts for small sample bias:

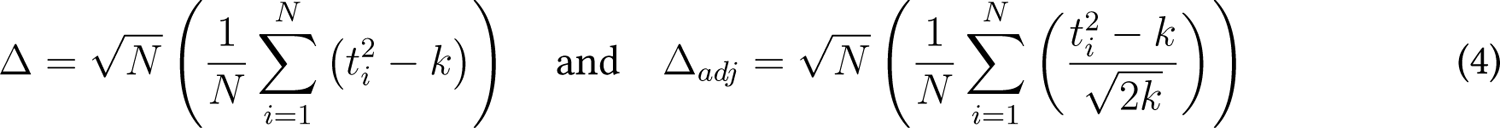

where: *N* is the number of cross-sectional units, *k* is the number of regressors,*t_i_* is the t-statistic of the *i*-th cross-sectional unit. These statistics test the null hypothesis of slope homogeneity against the alternative hypothesis of slope heterogeneity. A significant Δ or Δ*_adj_* indicates the presence of slope heterogeneity across the cross-sectional units. The results of slope heterogeneity estimated using Bersvendsen and Ditzen (2021) is given in table-11.

#### 2.2.3 Panel unit root test

For panels exhibiting clear cross-sectional dependence, Kapetanios, Pesaran, and Yamagata (2011) introduced the CIPS test, a second-generation panel unit root test. Later, Westerlund, Hosseink-ouchack, and Solberger (2016) derived its asymptotic properties, enhancing its robustness and making it superior to other panel unit root tests. The CIPS test accommodates cross-sectional dependence by incorporating weighted lag averages and differences for each panel unit, making it suitable for this study. The test is based on the cross-sectional augmented Dickey-Fuller (CADF) test and is formulated as follows:

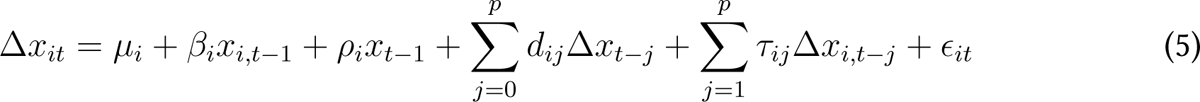

where *x_t−_*_1_ and Δ*x_t−j_* represent the cross-sectional lag averages and the first differences, with coefficients *ρ_i_* and *d_ij_*, respectively. *µ_i_* and *β_i_* denote the constants and drifts, while *τ_ij_* is the lead factor. The CIPS statistic, calculated using CADF statistics, is given by:

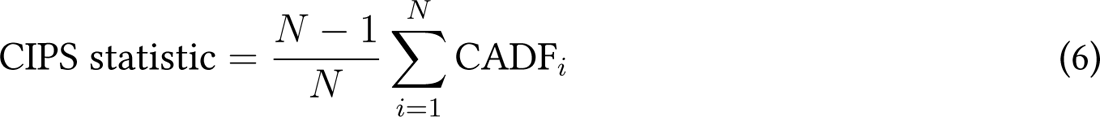

However, after finding slope heterogeneity we use Herwartz, Maxand, Raters, and Walle (2018) panel unit root test that is useful for heteroscedastic panels. The results of the panel unit root tests are presented in Table 8.

#### 2.2.4 Panel structural break test

I use Ditzen, Karavias, and Westerlund (2021)’s sequential test for multiple breaks at unknown breakpoints to find possible structural breaks in our data. for which I the sequential F-statistic to test for additional breaks:

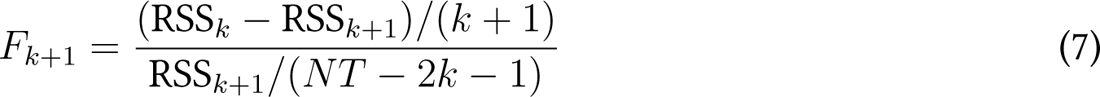

where *NT* is the total number of observations. Compare *F_k_*_+1_ to the critical value to determine if an additional break is significant. The test results are presented tn in table-10.

#### 2.2.5 Panel test for cointegration

The error correction panel cointegration test developed by Westerlund and Edgerton (2007) was utilized in this study, following the methodology outlined by Westerlund (2007). This test accommodates both within-unit and between-unit cross-sectional dependence by incorporating error correction terms in its computations. It examines two different null hypotheses: (1) the absence of cointegration within individual panel units, and (2) the absence of cointegration across all panel units. For the first null hypothesis, the group mean statistics *G_τ_*and *G_α_* are calculated using the estimated adjustment term *θ_i_* as follows:

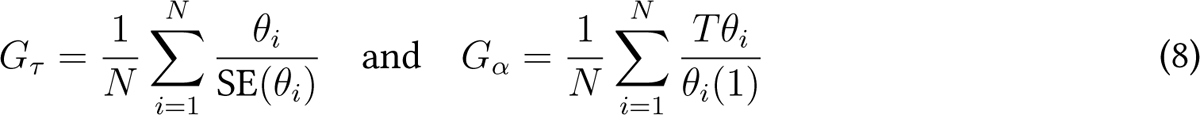

For the second null hypothesis, the group mean statistics are computed using these expressions:

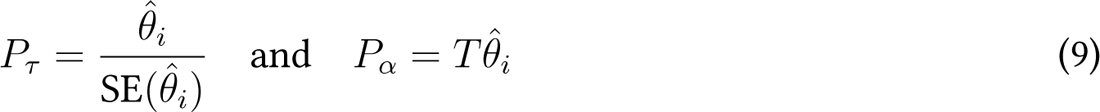

The results of Westerlund (2007)’s panel cointegration test are presented in table-9.

### 2.3 Panel estimators

To ensure the reliability of estimating the impact of NPIs on the incidence of COVID-19, this study employs state-of-the-art estimators. These modern estimators effectively account for cross-sectional dependence among variables by incorporating lagged averages to mitigate such effects. As we clearly see from Table 8, not all variables are stationary I(1), though they follow the I(1) process discussed in Herwartz et al. (2018) that accounts for heteroscedastic panels. Thus, the estimation framework used is Cross-Sectional Augmented Autoregressive Distributed Lags (CS-ARDL), which addresses this issue. Additionally, we utilize panel quantile and distribution estimators for several reasons. *Firstly,* because the effectiveness of NPIs may differ significantly for lower and higher quantiles of these outcomes, capturing potential disparities in response efficacy. *Secondly,* given the potential for heterogeneity in the distribution of COVID-19 data, quantile and distribution regressions provide a more comprehensive analysis compared to standard regression techniques. *Lastly,* these methods help validate the stability and consistency of our main regression results, offering a deeper understanding of how policy measures impact COVID-19 outcomes across different scenarios and population segments. Therefore, we present CS-ARDL and quantile & distribution regressions as individual estimators, as well as robustness checks for each other.

### 2.4 Panel cross-sectional augmented autoregressive distributed lags (CS-ARDL)

Chudik and Pesaran (2015) built a CS-ARDL estimator, which directly estimates long- and short-run regression coefficients between explanatory variables and variables of interest.

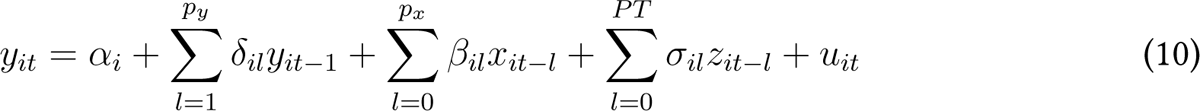

*^y^it* = *α i* where *i* = 1, 2*, …, N*, and 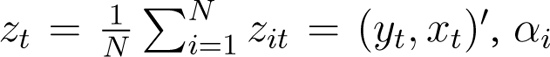 and *u_it_* are fixed effects and residuals, respectively. The model coefficients are computed using the following expression:

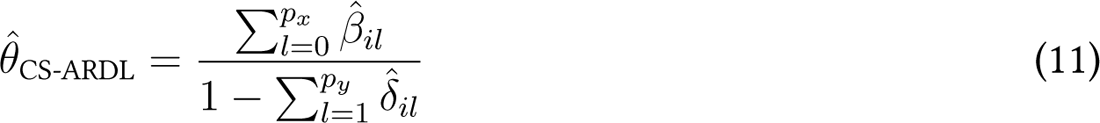

#### 2.4.1 Panel test for causalities

Dumitrescu and Hurlin (2012) introduced a causality test designed to ascertain the direction of causation between variables. This test is particularly suitable for extensive datasets and is known for producing reliable and robust outcomes even in the presence of cross-sectional dependence among variables, as highlighted by Fahimi, Saint Akadiri, Seraj, and Akadiri (2018). The causality test examines three main hypotheses: (1) Feedback or bidirectional causation, (2) Unidirectional causation, moving conservatively or growth-wise from one variable to another, and (3) The neutral hypothesis.

The mathematical representation of the test is given by:

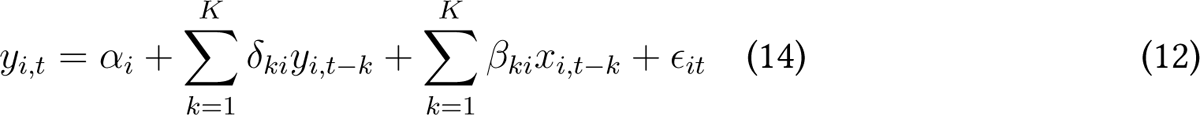

where *y* and *x* represent the variables under examination, *α* denotes the static impact, *δ* and *β* are autoregressive and reversion coefficients, respectively, and *K* denotes the optimal lag selected for all cross-sectional components.

The Wald statistic, critical for testing Granger non-causality across cross-sectional components, is computed as:

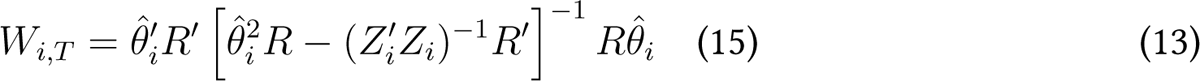

#### 2.4.2 Panel quantile and distribution regression

Panel quantile regression introduced by Koenker and Bassett Jr (1978) is the method of preference when we are interested in the effect of a policy on the distribution of an outcome. However, this panel quantile estimator estimates the *τ*-th conditional quantile *Q_τ_* (*y_it_|***x***_it_*) of the dependent variable *y_it_* given panel-specific independent variables **x***_it_*. The model formulates this quantile as:

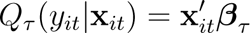

where ***β****_τ_* represents quantile-specific coefficients. Minimum Distance Estimation (MDE) suggested by Melly and Pons (n.d.) is employed to handle heterogeneity across panels and estimate these coefficients robustly. Figure-7 shows the graph of quantiles for each variable of the model to verify its heterogeneous behaviour concerning a normal distribution.

## 3 Results and discussion

### 3.1 Pretesting results

The pretesting results follow the *scheme of estimation procedures* depicted in Figure 2. **(1)** Findings from tests for cross-sectional dependence (Pesaran, 2021; Breusch & Pagan, 1980), as presented in Table 7, indicate that no null hypothesis of cross-sectional independence is rejected across different significance levels. Results from the cross-sectional dependence exponent test (Bailey et al., 2016) show that *α ≈* 1, indicating significant cross-sectional dependence, detailed in the last column of Table 7. Therefore, the results suggest the presence of cross-sectional dependence among variables. **(2)** Results from the slope heterogeneity test (Pesaran & Yamagata, 2008; Blomquist & Westerlund, 2013; Bersvendsen & Ditzen, 2021), detailed in Table 11, reveal statistically significant adjusted delta statistics for all three estimators, confirming the presence of slope heterogeneity. **(3)** Results from the CIPS panel unit root test (Pesaran, 2007), as presented in Table 8, reject the null hypothesis of panel unit root at levels with constant-trend for all variables except for LnECONOMIC. This hypothesis is also rejected at first difference with a trend for all variables. However, the unit root test by Herwartz et al. (2018), more suitable for heteroscedastic panels, shows that all variables are stationary at first differences, as shown in column 3 of Table 8. These findings imply first-order cointegration of all variables according to Herwartz et al. (2018), while Pesaran (2007) identifies variables as both I(0) and I(1). Therefore, Westerlund and Edgerton (2007) cointegration tests are appropriate to detect long-run relationships among variables, confirming CS-ARDL as the suitable estimation method. **(4)** Results from the structural breaks analysis are provided in Table 10. The test statistic values are below Bai & Perron’s critical values for structural break tests (Ditzen, 2021) at all significance levels, indicating no detected breaks in the data. **(5)** Cointegration findings by Westerlund and Edgerton (2007), presented in Table 9, reject the null hypothesis of no cointegration across the panel units, suggesting the presence of cointegration among all variables. These findings support long-term causal relationships among LnDEATHS, LnINFECTIONS, LnOVERALL, LnSTRINGENCY, LnCONTAINMENT, and LnECONOMIC across the 33 states and union territories of India from 2020 to 2022.

### 3.2 Estimator Results

#### 3.2.1 CS-ARDL Results

Findings related to short-term and long-term relationships between LnDEATHS and NPIs, as well as between LnINFECTIONS and NPIs, estimated from the CS-ARDL model are presented in Table 1 and Table 2, respectively.

**Table 1:** Results from CS-ARDL analysis with Deaths due to COVID-19 as dependent variables.

| *d.LnDEATHS* | *Overall* |  | *Stringency* |  | *Containment* |  | *Economic Support* |  |
| --- | --- | --- | --- | --- | --- | --- | --- | --- |
|  | (1) |  | (2) |  | (3) |  | (4) |  |
| *Short-term* |  |  |  |  |  |  |  |  |
| LnINFECTIONS | 0.680** | (0.210) | 0.686** | (0.203) | 0.681* | (0.212) | 0.659*** | (0.198) |
| L.LnINFECTIONS | 0.00457 | (0.0580) | 0.00363 | (0.0647) | 0.00766 | (0.0574) | -0.00150 | (0.0683) |
| LnOVERALL | -0.435*** | (0.0499) |  |  |  |  |  |  |
| LnECONOMIC |  |  |  |  |  |  | -0.221*** | (0.0386) |
| LnCONTAINMENT |  |  |  |  | -0.428*** | (0.0489) |  |  |
| LnSTRINGENCY |  |  | -0.331*** | (0.0365) |  |  |  |  |
| Constant | 0.817** | (0.195) | 0.801* | (0.198) | 0.806* | (0.195) | 0.491** | (0.129) |
| *Long-term* |  |  |  |  |  |  |  |  |
| Lr.LnINFECTIONS | -0.995** | (0.0580) | -0.996* | (0.0647) | -0.992* | (0.0574) | -1.002** | (0.0683) |
| Lr.LnOVERALL | -0.535*** | (0.0821) |  |  |  |  |  |  |
| Lr.LnECONOMIC |  |  |  |  |  |  | -0.361** | (0.122) |
| Lr.LnCONTAINMENT |  |  |  |  | -0.527*** | (0.0794) |  |  |
| Lr.LnSTRINGENCY |  |  | -0.440*** | (0.0768) |  |  |  |  |
| Lr.CONSTANT | 0.923** | (0.0656) | 0.956* | (0.0888) | 0.912* | (0.0640) | 0.608** | (0.0740) |
| *N* | 4950 |  | 4950 |  | 4950 |  | 4950 |  |
| *R*<sup>2</sup> | 0.520 |  | 0.526 |  | 0.528 |  | 0.516 |  |
| *Adj. R*<sup>2</sup> | 0.490 |  | 0.495 |  | 0.498 |  | 0.485 |  |
| *Standard errors in parentheses — \* $p<0.05$, \*\* $p<0.01$, \*\*\* $p_i0.001$* |  |  |  |  |  |  |  |  |

**Table 2:** Results from CS-ARDL analysis with infections of COVID-19 as dependent variables.

| *Dependent Variable*<br>*d.LnINFECTIONS* | *Overall* | *Stringency* | *Containment* | *Economic Support* |
| --- | --- | --- | --- | --- |
|  | (5) | (6) | (7) | (8) |
|  | *Short Run* |  |  |  |
| LnOVERALL | 0.00261 (0.0330) |  |  |  |
| LnECONOMIC |  |  |  | 0.000870 (0.0143) |
| LnCONTAINMENT |  |  | 0.00420 (0.0338) |  |
| LnSTRINGENCY |  | 0.00986 (0.0240) |  |  |
| CONSTANT | 0.0102 (0.126) | -0.00534 (0.119) | 0.00588 (0.134) | 0.0110 (0.0789) |
|  | *Long Run* |  |  |  |
| lr_LnOVERALL | -0.997*** (0.0330) |  |  |  |
| lr_LnSTRINGENCY |  | -0.990*** (0.0240) |  |  |
| lr_LnCONTAINMENT |  |  | -0.996*** (0.0338) |  |
| lr_LnECONOMIC |  |  |  | -0.999*** (0.0143) |
| lr__CONSTANT | 0.0106** (0.000339) | -0.00549* (0.000127) | 0.00612* (0.000210) | 0.0111** (0.000142) |
| *N* | 4983 | 4983 | 4983 | 4983 |
| *R*<sup>2</sup> | 0.919 | 0.930 | 0.921 | 0.912 |
| *Adj. R*<sup>2</sup> | 0.915 | 0.927 | 0.917 | 0.908 |
| *Standard errors in parentheses — \* $p < 0.05$, \*\* $p < 0.01$, \*\*\* $p < 0.001$* |  |  |  |  |

In the short-term analysis in table-1, the coefficients indicate a significant relationship between variousNPIs and the logarithmic change in COVID-19 related deaths (d.LnDEATHS). For the overall government response (Column 1), the infection rate (LnINFECTIONS) has a significant positive coefficient of 0.680 (p*<*0.01), indicating that higher infection rates are associated with increased deaths. Similarly, the coefficients for infection rates in columns 2, 3, and 4 (0.686, 0.681, and 0.659 respectively, all p*<*0.01) show a consistent positive impact. This finding is obvious as infections and deaths are highly expected to move together (see Malki et al. (2020)) in the short-term; however in the longer-term with pharmaceutical and non-pharmaceutical interventions in place this relation may get reversed. The stringent measures (Column 2), containment measures (Column 3), and economic support measures (Column 4) have negative coefficients (−0.331, −0.428, and −0.221, all p*<*0.001), suggesting that stricter measures effectively reduce the number of deaths in the short term. These results highlight the crucial role of immediate and stringent government interventions in controlling the death toll during the pandemic’s early stages, which further confirmed by the −0.435 (p*<*0.001) coefficient of overall government support that combines the impact of all the three NPIs.

In the long-term analysis in table-1, the results continue to emphasize the importance of infection rates and NPIs. The coefficients for the infection rates (Lr LnINFECTIONS) remain highly significant and negative across all columns (−0.995, −0.996, −0.992, and −1.002, all p*<*0.001), indicating that the infection rates have a profound negative association with deaths over a prolonged period. This is primarily due to studies such as Stein et al. (2023), which have identified a significant reduction in the risk of COVID-19 infection among individuals with a previous SARS-CoV-2 infection compared to those without prior infection. Consequently, as more people get infected over time, the long-term susceptibility to further infections and deaths decreases. The coefficients for overall government response (Lr LnOVERALL) and specific measures like economic support (Lr LnECONOMIC), containment (Lr LnCONTAINMENT), and stringency (Lr LnSTRINGENCY) also show significant negative values (−0.535, −0.361, −0.527, and −0.440, respectively, all p*<*0.01). These results suggest that continuous and consistent application of NPIs significantly mitigates the mortality rate in the long run. These findings underscore the long-term benefits of maintaining rigorous public health measures and support systems to manage and reduce the impact of the pandemic effectively.

The interpretation of COVID-19 infection analyses requires careful consideration due to several factors. While infections are confirmed through testing, their reliability as an indicator of COVID-19 incidence is compromised by issues such as misreporting and variability in symptom presentation. This makes infections a less dependable metric compared to deaths. Moreover, short-term analyses focusing on infections often yield non-significant results. This is primarily because pharmaceutical interventions, particularly vaccinations, have a profound impact on infection rates, overshadowing the immediate effectiveness of non-pharmaceutical interventions (NPIs) like social distancing and mask mandates (Tenforde et al., 2021; Peters, Raymer, Pal, & Ambardekar, 2022). Additionally, the possibility of individuals being repeatedly infected further undermines the short-term efficacy of NPIs in controlling transmission. Despite these limitations associated with COVID-19 infections, focusing exclusively on deaths does not adequately capture the full impact of the pandemic. Infections have had far-reaching consequences, profoundly affecting physical health and social well-being globally. Therefore, while acknowledging the challenges in interpreting infection data, it remains crucial to consider both infections and deaths to comprehensively assess the public health impact of COVID-19. In the short-term analysis of the impact of various government measures on the logarithmic change in infections (d.LnINFECTIONS), the coefficients for all categories—overall government response, stringency measures, containment measures, and economic support measures—are statistically insignificant. Specifically, the coefficients for overall response (0.00261), stringency (0.00986), containment (0.00420), and economic support (0.000870) all fail to reach statistical significance, suggesting that these measures do not have an immediate effect on the infection rates in the short-run. This insignificance in the short term could be attributed to several factors as discussed in the previous paragraph. For one, there may be a delay between the implementation of policy measures and their observable effects on infection rates. Additionally, short-term infection rates can be influenced by various external factors, including public compliance and adherence fatigue (Petherick et al., 2021), variations in testing rates, and reporting delays, which may dilute the immediate impact of governmental interventions in the short-term.

In contrast, the long-term analysis reveals highly significant negative coefficients for all NPIs, underscoring their effectiveness in controlling infections over a sustained period. The coefficients for the overall government response (lr LnOVERALL) and specific measures like stringency (lr - LnSTRINGENCY), containment (lr LnCONTAINMENT), and economic support (lr LnECONOMIC) are all close to −1 (−0.997, −0.990, −0.996, and −0.999, respectively, all p*<*0.001). These results indicate that sustained implementation of these measures significantly reduces infection rates in the long run. These findings highlight the critical importance of maintaining robust public health measures over time to effectively curb the spread of the virus, as immediate effects may not always be apparent, but the long-term benefits are substantial and measurable.

#### 3.2.2 Causalities results

The dependent variables, deaths and infections due to COVID-19, may exhibit a bi-directional relationship with intervention measures (NPIs), implying that these measures both influence deaths, infection and are influenced by deaths. The table-3 presents the results from the JKS non-causality test (Juodis, Karavias, Sarafidis, 2021) to examine the bidirectional relationships between various government response measures, infection rates, and death rates during the COVID-19 pandemic. The Wald statistics and coefficients for each causal relationship provide insight into the direction and nature of these effects. The overall government response exhibits a significant negative causal effect on death rates (Wald Stat: 220.68, Coefficient: −0.926, p*<*0.001), indicating that stronger overall government interventions lead to a reduction in death rates. Conversely, there is a significant positive causal effect of death rates on overall government response (Wald Stat: 132.26, Coefficient: 0.0324, p*<*0.001), suggesting that higher death rates prompt an increase in government interventions.

Stringency measures have a significant negative impact on death rates (Wald Stat: 210.80, Coefficient: −0.0326, p*<*0.001), implying that stringent policies effectively reduce death rates. However, the feedback from death rates to stringency measures is positive and significant (Wald Stat: 487.37, Coefficient: 0.0642, p*<*0.001), indicating that increasing death rates result in stricter policies.

Containment measures significantly reduce death rates (Wald Stat: 285.66, Coefficient: −1.08, p*<*0.001), while rising death rates significantly increase containment measures (Wald Stat: 61.0983, Coefficient: 0.022, p*<*0.001). This bidirectional causality underscores the reactive nature of containment measures to death rates. Economic support measures show a strong negative causal effect on death rates (Wald Stat: 212.56, Coefficient: −0.775, p*<*0.001). Conversely, higher death rates significantly increase economic support measures (Wald Stat: 620.63, Coefficient: 0.12, p*<*0.001), highlighting the importance of economic interventions in response to rising death rates. Government responses also affect infection rates. The overall government response has a negative causal effect on infections (Wald Stat: 35.24, Coefficient: −0.48, p*<*0.001), while infections positively impact the overall response (Wald Stat: 778.98, Coefficient: 0.0689, p*<*0.001). Stringency measures, although having an insignificant effect on infections (Wald Stat: 360.58, Coefficient: −0.0018), are significantly influenced by infection rates (Wald Stat: 302.45, Coefficient: 0.13, p*<*0.001). Containment measures reduce infections (Wald Stat: 49.47, Coefficient: −0.586, p*<*0.001), and infections lead to increased containment (Wald Stat: 495.99, Coefficient: 0.0544, p*<*0.001). Economic support measures also reduce infections (Wald Stat: 44.31, Coefficient: −0.277, p*<*0.001), with infections significantly increasing economic support measures (Wald Stat: 390.56, Coefficient: 0.249, p*<*0.001). There is a strong negative causal effect of infections on deaths (Wald Stat: 170.65, Coefficient: 1.89, p*<*0.001), while deaths also negatively impact infections (Wald Stat: 747.95, Coefficient: −1.803, p*<*0.001).

#### 3.2.3 Quantile regression results

Quantile regression results for LnDEATHS as the dependent variable are presented in Tables 3 to 5, while results for LnINFECTIONS are found in Tables 6 to 8 in the supplementary materials. Summary statistics of the variable quantiles are provided in Table 2. Additionally, Figure 4 and figure 5 display the quantile plots corresponding to these regressions with LnDEATHS and LnINFECTIONS as the dependent variables, respectively.

**Figure 4:**
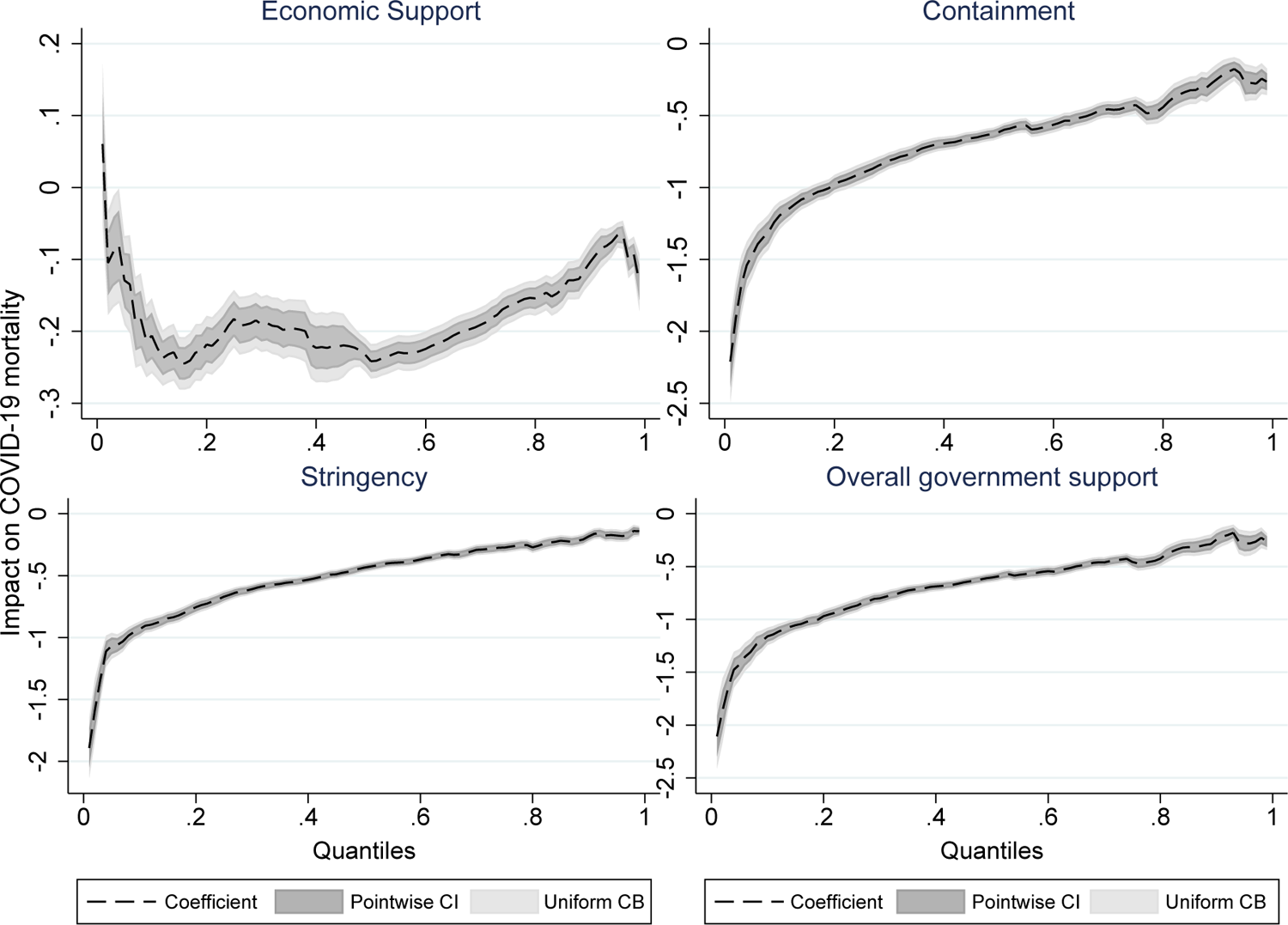
Quantile regression plots of NPIs in reducing deaths due to COVID-19.

**Figure 5:**
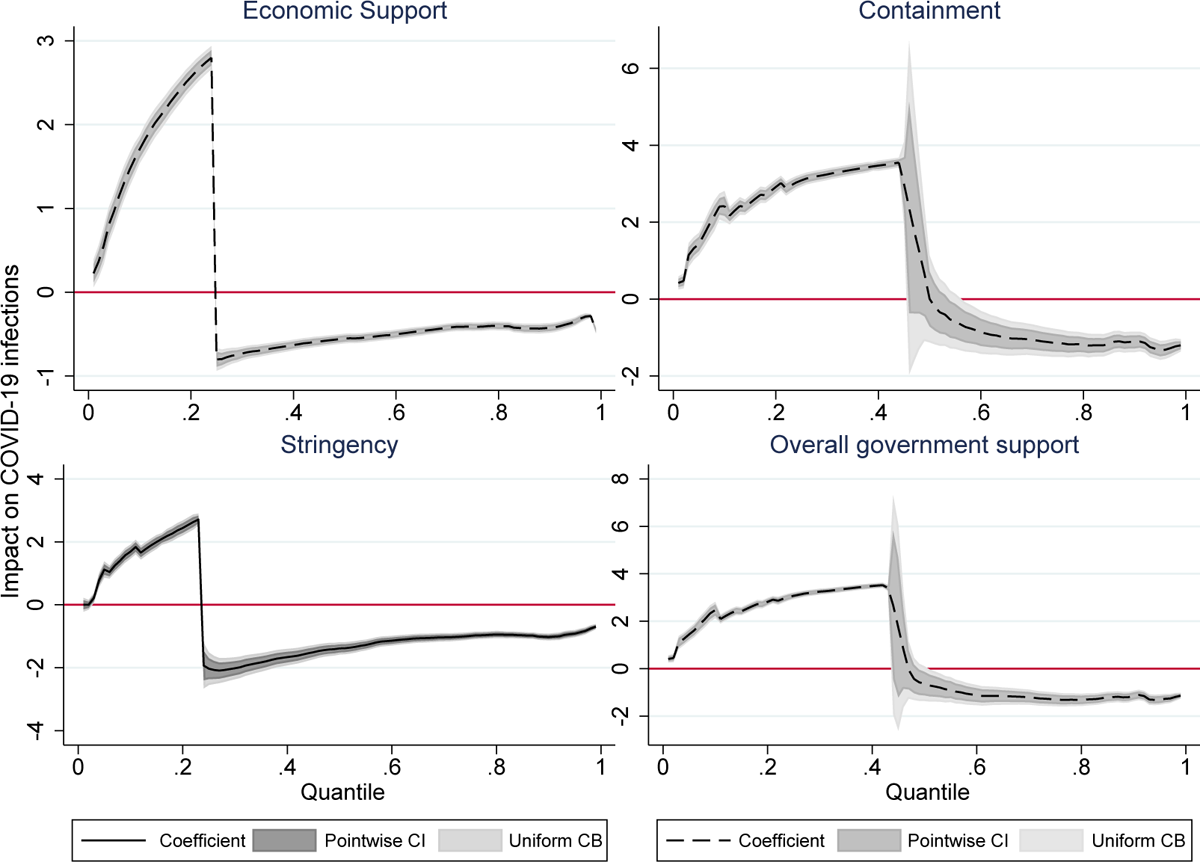
Quantile regression plots of NPIs in reducing infections of COVID-19.

**Table 3:**
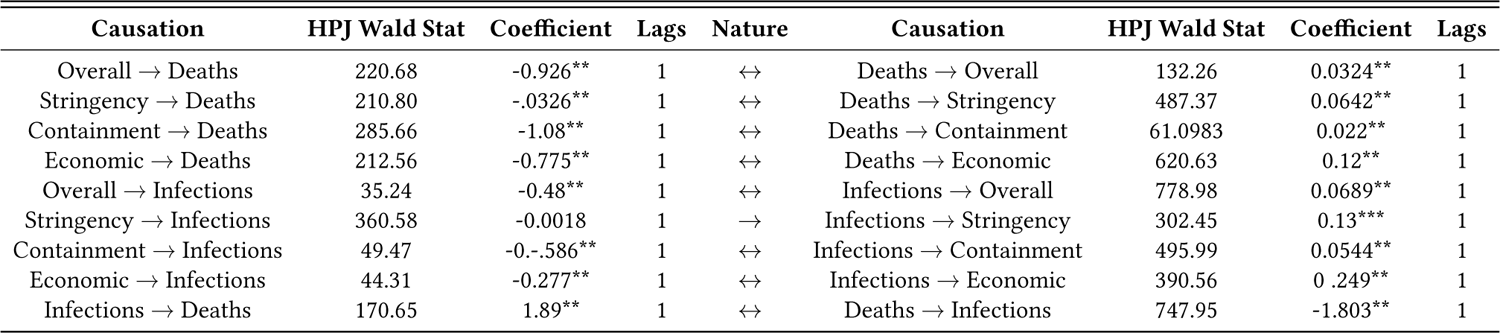
JKS Non-causality test (Juodis, Karavias, Sarafidis, 2021)

| Causation | HPJ Wald Stat | Coefficient | Lags | Nature | Causation | HPJ Wald Stat | Coefficient | Lags |
| --- | --- | --- | --- | --- | --- | --- | --- | --- |
| Overall → Deaths | 220.68 | -0.926** | 1 | ↔ | Deaths → Overall | 132.26 | 0.0324** | 1 |
| Stringency → Deaths | 210.80 | -0.0326** | 1 | ↔ | Deaths → Stringency | 487.37 | 0.0642** | 1 |
| Containment → Deaths | 285.66 | -1.08** | 1 | ↔ | Deaths → Containment | 61.0983 | 0.022** | 1 |
| Economic → Deaths | 212.56 | -0.775** | 1 | ↔ | Deaths → Economic | 620.63 | 0.12** | 1 |
| Overall → Infections | 35.24 | -0.48** | 1 | ↔ | Infections → Overall | 778.98 | 0.0689** | 1 |
| Stringency → Infections | 360.58 | -0.0018 | 1 | → | Infections → Stringency | 302.45 | 0.13*** | 1 |
| Containment → Infections | 49.47 | -0.586** | 1 | ↔ | Infections → Containment | 495.99 | 0.0544** | 1 |
| Economic → Infections | 44.31 | -0.277** | 1 | ↔ | Infections → Economic | 390.56 | 0.249** | 1 |
| Infections → Deaths | 170.65 | 1.89** | 1 | ↔ | Deaths → Infections | 747.95 | -1.803** | 1 |

The quantile regression results reveal significant insights into how different policy measures impact COVID-19 related deaths across various quantiles. Across all quantiles (Q1 to Q10), the logarithm of infections (LnINFECTIONS) consistently shows a strong positive relationship with the logarithm of deaths (LnDEATHS), indicating that higher infection rates are strongly associated with higher death rates confirming the findings in table-1. The coefficients for LnINFECTIONS gradually decrease from Q1 (0.973) to Q10 (0.756), suggesting a slightly diminishing effect of infections on deaths as the severity of the death rate increases, which can be mainly due to the fact that pharmaceutical interventions come into play. The overall government response (LnOVERALL) has a significant negative impact on death rates across all quantiles, with the strength of this negative relationship diminishing slightly from Q1 (−1.428) to Q10 (−0.280). This indicates that comprehensive government responses are effective in reducing death rates, especially in lower quantiles where the death rates are not extremely high. Similarly, stringent measures (LnSTRINGENCY), containment measures (LnCONTAINMENT), and economic support measures (LnECONOMIC) all show a consistent negative relationship with death rates, suggesting that these interventions are crucial in mitigating the impact of the pandemic. The coefficients for these measures also generally decrease in absolute value across higher quantiles, implying that their effectiveness in reducing death rates is more pronounced at lower death rates.

The quantile regression analysis reveals significant relationships between COVID-19 infection rates (‘d.LnINFECTIONS‘) and various governmental response measures across different quantiles. For instance, in the lowest quantile (Q1), an increase in the overall government response index (‘LnOVERALL‘) by 1 unit is associated with an average increase in ‘d.LnINFECTIONS‘ by 1.447 (SE = 0.0881). Similarly, stricter stringency measures (‘LnSTRINGENCY‘) show a significant effect across quantiles, such as a coefficient of 1.119 (SE = 0.0918) in Q1. Containment measures (‘LnCONTAINMENT‘) also exhibit notable impacts, with coefficients like 1.434 (SE = 0.0905) in Q1, indicating their positive association with infection rates. Economic support measures (‘LnECONOMIC‘) display consistent positive coefficients across all quantiles, such as 0.967 (SE = 0.0687) in Q1, suggesting that higher economic support correlates with increased infection rates. These findings underscore the fact that is established in table-2 that in the short-term NPIs are largely ineffective in controlling infection. However, as also seen in figure-5 after Q2 for economic support, and stringency and after Q4 for containment and overall government response, NPIs start to become more effective. It is important to note that, as over time more and more NPIs come into place the NPI index scores increase, so higher quantiles necessarily indicate a later time period and vice versa.

## 4 Policy Implications & Limitations of the study

The results from the previous section highlight several key policy suggestions that can effectively guide responses to pandemics or endemics, especially when pharmaceutical interventions are delayed and innovation has a gestation period. Given global inequalities that obstruct the reach of timely pharmaceutical interventions (Sparke & Levy, 2022; Hakobyan, Rawlings, & Yao, 2022; Bayati, Noroozi, Ghanbari-Jahromi, & Jalali, 2022), it is crucial to implement appropriate non-pharmaceutical interventions (NPIs) to control the incidence of such pandemics. The policy suggestions are as follows:

1. In the short term, all NPIs show a significant and negative association with COVID-19 incidences of deaths and infections, indicating their effectiveness in controlling the endemic. Additionally, in densely populated countries like India, policies of containment and stringency have been observed to be particularly effective. Particularly, as given the vaccine gestation period stringent measures are most effective as demonstrated in quantile regression where higher stringency values have demonstrated better effect. Moreover as reinstated in the causality analysis, more infection and deaths lead to further stringent and stricter NPIs that intern further reduce the COVID-19 incidence.
2. Taking death into consideration, overall NPI impact is impressive, however the economic support signals further emphasis. Though studies like Varshney, Kumar, Mishra, Rashid, and Joshi (2021) find that, government transfer packages during the pandemic were significant in alleviating credit constraints and played a role in furthering agricultural investments, there has been several doubts casted over the financial aids data used in these studies. Moreover the fact that India didn’t have a robust direct transfer plan during the COVID-19, when considered with the scant spending by the government when compared to other nations, caste further doubt about what could have been the impact of economic support had there been support considerably. This warrants for significant economic support push in such situations in India.
3. Several factors like political trust (Ji, Jiang, & Zhang, 2024) also influence the impact of government response to COVID-19 as they impact the effective implementation of NPIs, which are often rumoured to be ineffective in the absence of pharmaceutical interventions. Moreover, the fact that interventions like economic support also help in making the economy bouncing back easily by means of gained consumer confidence (Gholipour, Tajaddini, & Farzanegan, 2023), which indicates towards multifaceted benefits of NPIs during the pandemic situations.
4. The major insight that emerges from the causality analysis is that NPIs have been observed to be informed with the changes warrant in the health landscape during the pandemic. The fact that the causal direction from NPIs to COVID-19 incidence are negative and vice versa, suggests that higher deaths have led to stricter interventions and stricter interventions have further resulted in curbing deaths and infections.
5. Crucial interpretation of quantile regression suggests two major implications. *First,* not all levels of NPIs are equally effective conditioned on several other factors, thus monitoring of interventions in places is duly warranted and flexibility in tightening and relaxing interventions as the need may arise is the key to better management of pandemics. *Second,* considering human movement to an extent inevitable for survival, the long-term effect is delayed though effective at last. However, it is further confirmed that, deaths are controlled using NPIs more effectively than infections.
6. The fact that this study confirms long-run effectiveness of NPIs in curbing both infections and deaths due to COVID-19 the major implication presents NPIs as unavoidable interventions during pandemic irrespective of the pharmaceutical interventions like vaccines. Certainly, their effectiveness in the long-run is entangled with that of the vaccines, yet it is established by the quantile regression that, in controlling infections the NPIs were initially ineffective however, they have proven to be effective in the long-run. Conversely, they are even better in controlling deaths.

Though the study confirms several intuitive validations of NPIs during pandemic and contests against the role of them compared to pharmaceutical interventions, this study does have several *limitations*. *First,* while it established robust associations, it does not establish any causal relationships that isolates the influence of other important covariates. Thus, the observed correlations between NPIs and COVID-19 incidences cannot be interpreted as definitive evidence of causation. *Second,* the study is constrained by its inability to control for all variables that impact COVID-19 incidence. There are numerous human-related, governmental, social, biological, and environmental factors at play, making it challenging to account for all of them in a single analysis. This limitation means that some confounding factors may influence the results. *Third,* a significant drawback is the study’s inability to isolate the influence of vaccines on controlling deaths and infections. The rollout of vaccines during the study period adds another layer of complexity, as it is difficult to disentangle the effects of NPIs from those of vaccination efforts.

## 5 Conclusion

In summary, this paper highlights the significant impact of non-pharmaceutical interventions (NPIs) on controlling COVID-19 deaths and infections. Our analysis reveals that NPIs, such as containment and stringency measures, have been effective, particularly in densely populated regions like India. The findings underscore the importance of implementing comprehensive NPIs to manage pandemics, especially in the absence of timely pharmaceutical solutions. Furthermore, the paper emphasizes the multifaceted benefits of NPIs, including their role in boosting consumer confidence and aiding economic recovery. Overall, the research provides valuable insights for policymakers and public health officials in crafting effective strategies for current and future pandemics.

## Data Availability

All data produced are available online at OXCGRT Database

https://www.bsg.ox.ac.uk/research/covid-19-government-response-tracker

## Appendix

### A.1 OxCGRT indices and variables of the study

**Table 4:**
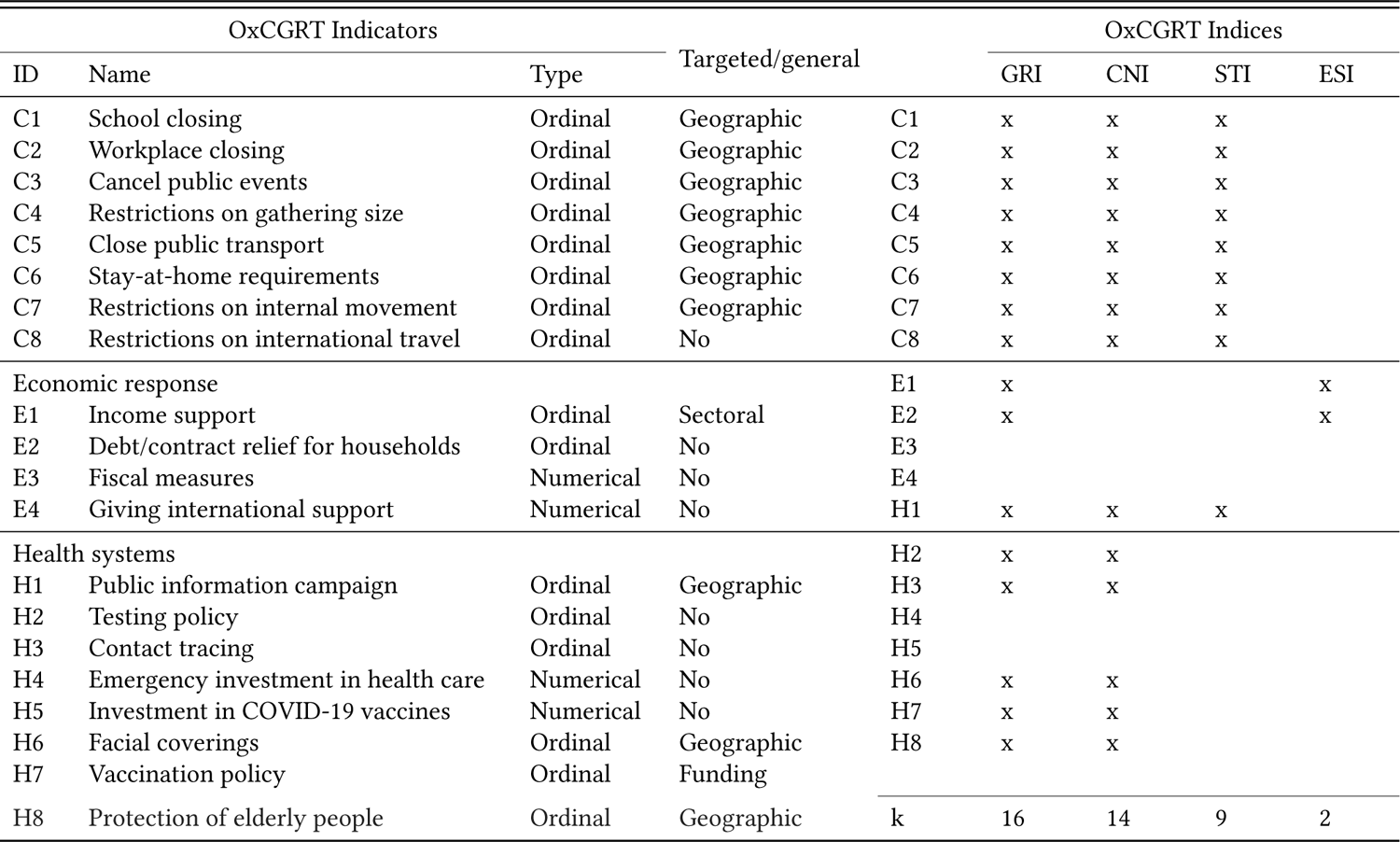
OxCGRT sub-indices and index variables of the study.

**Table 5:**
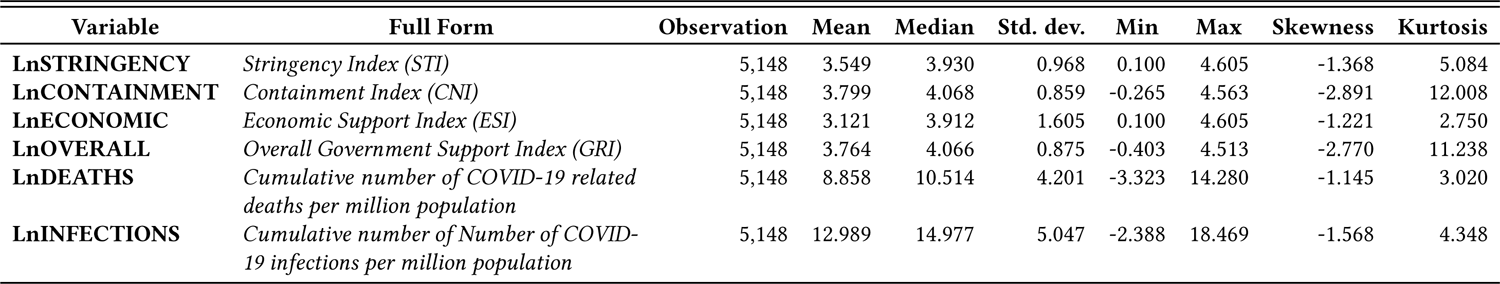
Descriptive statistics of Variables.

| Variable | Full Form | Observation | Mean | Median | Std. dev. | Min | Max | Skewness | Kurtosis |
| --- | --- | --- | --- | --- | --- | --- | --- | --- | --- |
| **LnSTRINGENCY** | *Stringency Index (STI)* | 5,148 | 3.549 | 3.930 | 0.968 | 0.100 | 4.605 | -1.368 | 5.084 |
| **LnCONTAINMENT** | *Containment Index (CNI)* | 5,148 | 3.799 | 4.068 | 0.859 | -0.265 | 4.563 | -2.891 | 12.008 |
| **LnECONOMIC** | *Economic Support Index (ESI)* | 5,148 | 3.121 | 3.912 | 1.605 | 0.100 | 4.605 | -1.221 | 2.750 |
| **LnOVERALL** | *Overall Government Support Index (GRI)* | 5,148 | 3.764 | 4.066 | 0.875 | -0.403 | 4.513 | -2.770 | 11.238 |
| **LnDEATHS** | *Cumulative number of COVID-19 related deaths per million population* | 5,148 | 8.858 | 10.514 | 4.201 | -3.323 | 14.280 | -1.145 | 3.020 |
| **LnINFECTIONS** | *Cumulative number of Number of COVID-19 infections per million population* | 5,148 | 12.989 | 14.977 | 5.047 | -2.388 | 18.469 | -1.568 | 4.348 |

**Table 6:**
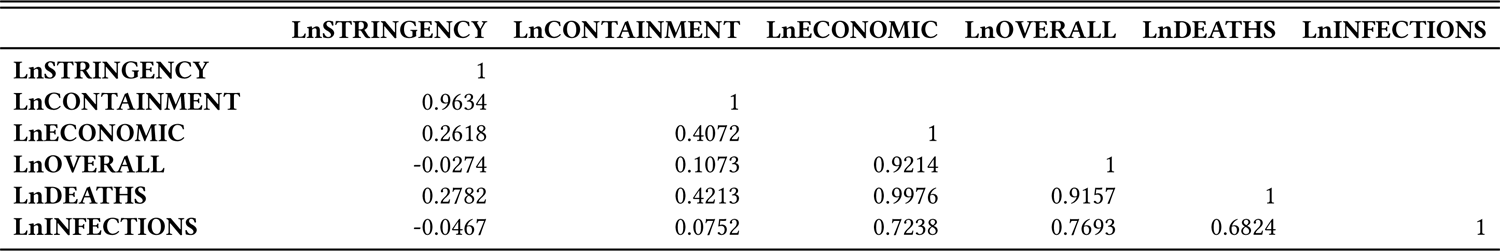
The Matrix of correlation.

**Table 7:** Cross - Sectional Dependence Results.

| Variable | BPLM | PLM | PCD | $\alpha$ |
| --- | --- | --- | --- | --- |
| **LnSTRINGENCY** | 41743.141** | 196.791* | 271.13** | 1.00098 |
| **LnCONTAINMENT** | 42921.748** | 184.673* | 279.73** | 1.0009 |
| **LnECONOMIC** | 44841.876** | 171.918* | 263.98** | 1.000919 |
| **LnOVERALL** | 42763.034** | 182.985* | 280.67** | 1.00091 |
| **LnDEATHS** | 18351.199** | 99.624* | 279.44** | 1.0009 |
| **LnINFECTIONS** | 19485.788** | 114.406* | 284.36** | 1.000 |

Alpha is a cross-sectional dependence exponent estimate that tests the level of identified cross-sectional dependence in the variable. With this test, if $\alpha \approx 1$ implies strong cross-sectional dependence. BPLM: Breusch Pagan Lagrange Multiplier, PLM: Pesaran Lagrange Multiplier, PCD: Pesaran CD.

**Table 8:** Results from panel unit root analysis.

| Variable at Level | CIPS - Pesaran (2007) | Herwartz et al. (2018) |
| --- | --- | --- |
| **LnSTRINGENCY** | -2.807*** | 1.6869 |
| **LnCONTAINMENT** | -2.967*** | 1.3531 |
| **LnECONOMIC** | -1.729 | 1.5807 |
| **LnOVERALL** | -2.947*** | 1.1112 |
| **LnDEATHS** | -2.355*** | 1.5807 |
| **LnINFECTIONS** | -3.063*** | 2.0150 |
| Variable at 1st Difference with Trend | CIPS - Pesaran (2007) | Herwartz et al. (2018) |
| **LnSTRINGENCY** | -6.055** | -2.3741** |
| **LnCONTAINMENT** | -6.166** | -1.8344* |
| **LnECONOMIC** | -6.393** | -1.5667* |
| **LnOVERALL** | -6.124** | -1.7357** |
| **LnDEATHS** | -5.034** | -2.9020** |
| **LnINFECTIONS** | -5.500** | -2.9890** |

**Table 9:** Panel Cointegration results.

| Statistic | Value | Z-value | P-value |
| --- | --- | --- | --- |
| **Gt** | -4.950*** | -13.888 | 0.00 |
| **Ga** | -87.907*** | -50.825 | 0.00 |
| **Pt** | -17.404*** | -3.490 | 0.00 |
| **Pa** | -56.851*** | -31.391 | 0.00 |

**Table 10:** Sequential test for multiple breaks at unknown breakpoints (Ditzen, Karavias & Westerlund. 2021)

| Bai & Perron Critical Values |  |  |  |  |
| --- | --- | --- | --- | --- |
|  | Test Statistic | 1% Critical Value | 5% Critical Value | 10% Critical Value |
| **F(10)** | 4.01 | 12.29 | 8.58 | 7.04 |
| **F(21)** | 2.33 | 13.89 | 10.13 | 8.51 |
| **F(32)** | 0.78 | 14.8 | 11.14 | 9.41 |
| **F(43)** | 0.26 | 15.28 | 11.83 | 10.04 |
| **F(54)** | 0.43 | 15.76 | 12.25 | 10.58 |
| **Detected number of breaks:** |  | - | - | - |

**Table 11:**
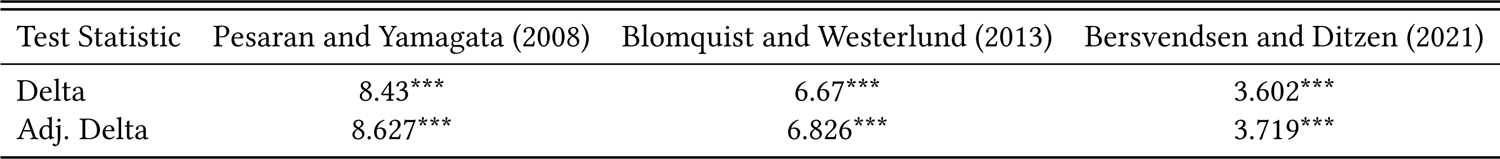
Tests’ results from slope heterogeneity analysis.

| Test Statistic | Pesaran and Yamagata (2008) | Blomquist and Westerlund (2013) | Bersvendsen and Ditzen (2021) |
| --- | --- | --- | --- |
| Delta | 8.43*** | 6.67*** | 3.602*** |
| Adj. Delta | 8.627*** | 6.826*** | 3.719*** |

### A.2 Diagnostic Test Results

**Figure 6:**
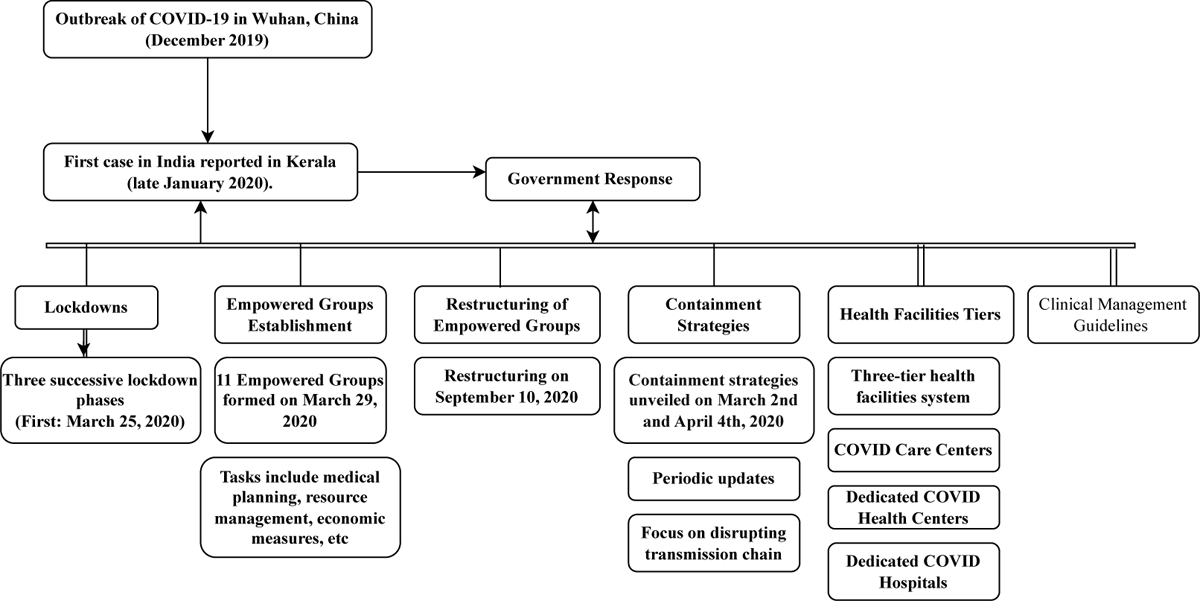
The framework of Indian management of COVID-19 pandemic (a) This figure depicts the different work-action measures taken as a government response since the first COVID case in India in January 2020. It shows, along with a phased implemented (and withdrawn) lockdown, there were containment strategies in place. For better functioning, 11 empowered groups were formed and further restructured to implement pharmaceutical and non-pharmaceutical guidelines.

**Figure 7:**
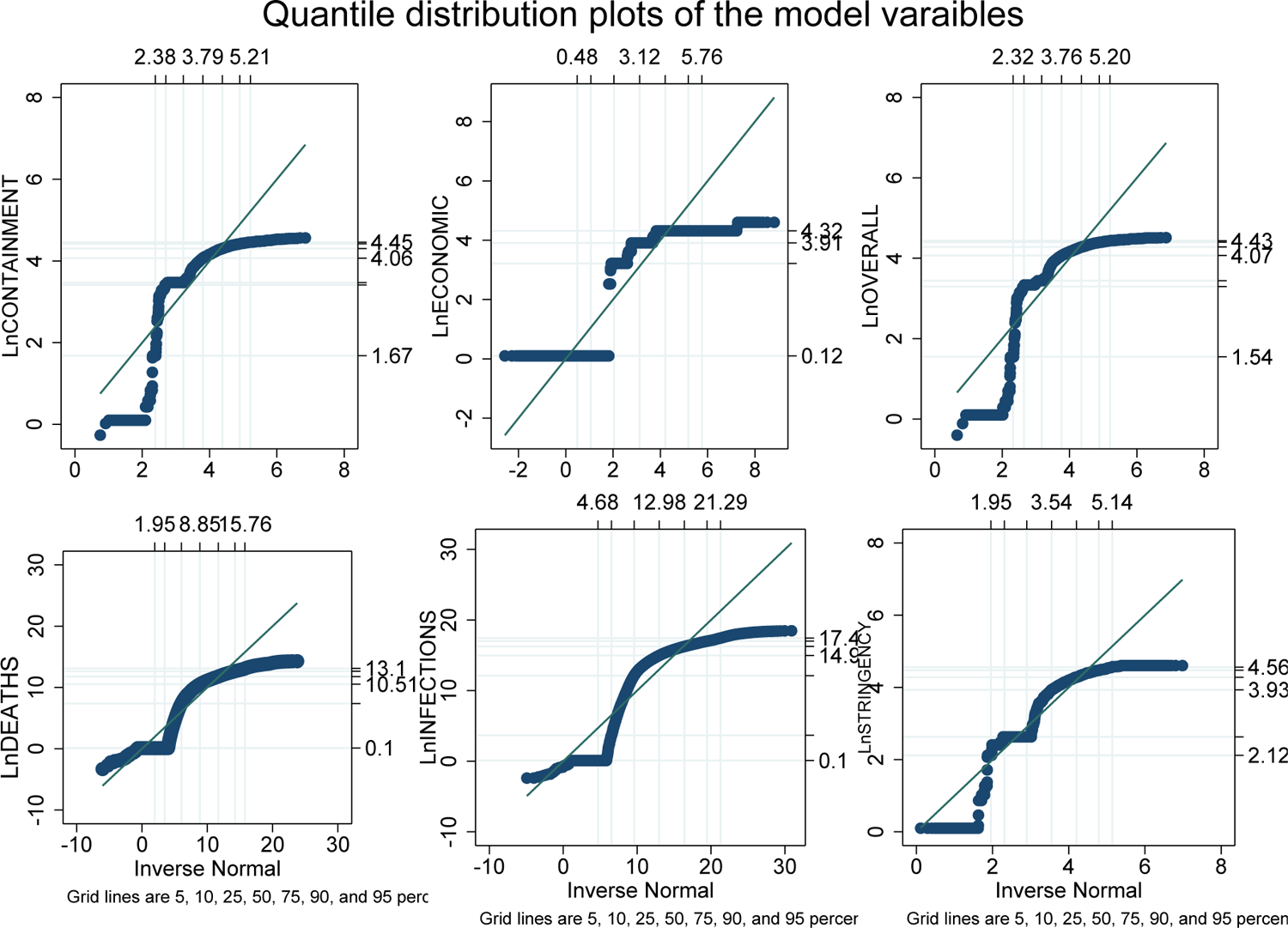
Quantile distribution plots of all variables.

## Footnotes

1 With a projected real GDP growth rate of 6.3%, compared to China (5%), USA (2.1%), U.K. (0.5%). For more details, see World Economic Outlook (October 2023).

2 Currently, as of 2022, India has 29 states and 8 Union Territories in India (Taylor & Shrimankar, 2024), however we take only 33 states and union territories as the data pertains to these sub-national entities only.

3 The first COVID-19 case was confirmed in India; (See Andrews et al. (2020) for more).

4 See Table-4

## References

Andrews, M., Areekal, B., Rajesh, K., Krishnan, J., Suryakala, R., Krishnan, B., … Santhosh, P. (2020). First confirmed case of covid-19 infection in india: A case report. The Indian journal of medical research, 151(5), 490.

Bailey, N., Kapetanios, G., & Pesaran, M. H. (2016). Exponent of cross-sectional dependence: Estimation and inference. Journal of Applied Econometrics, 31(6), 929–960.

Bayati, M., Noroozi, R., Ghanbari-Jahromi, M., & Jalali, F. S. (2022). Inequality in the distribution of covid-19 vaccine: a systematic review. International journal for equity in health, 21(1), 122.

Bersvendsen, T., & Ditzen, J. (2021). Testing for slope heterogeneity in stata. The Stata Journal, 21(1), 51–80.

Björk, J., Mattisson, K., & Ahlbom, A. (2021). Impact of winter holiday and government responses on mortality in europe during the first wave of the covid-19 pandemic. European journal of public health, 31(2), 272–277.

Bjørnskov, C. (2021). Did lockdown work? an economist’s cross-country comparison. CESifo Economic Studies, 67 (3), 318–331.

Blanco, F., Emrullahu, D., & Soto, R. (2020). Do coronavirus containment measures work? worldwide evidence. The World Bank.

Blomquist, J., & Westerlund, J. (2013). Testing slope homogeneity in large panels with serial correlation. Economics Letters, 121(3), 374–378.

Bollyky, T. J., Hulland, E. N., Barber, R. M., Collins, J. K., Kiernan, S., Moses, M., … others (2022). Pandemic preparedness and covid-19: an exploratory analysis of infection and fatality rates, and contextual factors associated with preparedness in 177 countries, from jan 1, 2020, to sept 30, 2021. The Lancet, 399(10334), 1489–1512.

Bonardi, J.-P., Gallea, Q., Kalanoski, D., & Lalive, R. (2020). Fast and local: How lockdown policies affect the spread and severity of covid-19. Covid Economics, 23, 325–351.

Bongaerts, D., Mazzola, F., & Wagner, W. (2021). Closed for business: The mortality impact of business closures during the covid-19 pandemic. PloS one, 16(5), e0251373.

Brauner, J. M., Mindermann, S., Sharma, M., Johnston, D., Salvatier, J., Gavenčiak, T., … others (2021). Inferring the effectiveness of government interventions against covid-19. Science, 371(6531), eabd9338.

Breusch, T. S., & Pagan, A. R. (1980). The lagrange multiplier test and its applications to model specification in econometrics. The review of economic studies, 47 (1), 239–253.

Chaturvedi, S., Porter, J., Pillai, G. K. G., Abraham, L., Shankar, D., & Patwardhan, B. (2023). India and its pluralistic health system–a new philosophy for universal health coverage. The Lancet Regional Health-Southeast Asia, 10.

Chernozhukov, V., Kasahara, H., & Schrimpf, P. (2021). Causal impact of masks, policies, behavior on early covid-19 pandemic in the us. Journal of econometrics, 220(1), 23–62.

Chudik, A., & Pesaran, M. H. (2015). Common correlated effects estimation of heterogeneous dynamic panel data models with weakly exogenous regressors. Journal of econometrics, 188(2), 393–420.

Chudik, A., Pesaran, M. H., & Tosetti, E. (2011). Weak and strong cross-section dependence and estimation of large panels. Oxford University Press Oxford, UK.

Ditzen, J. (2021). Estimating long-run effects and the exponent of cross-sectional dependence: An update to xtdcce2. The Stata Journal, 21(3), 687–707.

Ditzen, J., Karavias, Y., & Westerlund, J. (2021). Testing and estimating structural breaks in time series and panel data in stata. arXiv preprint arXiv:2110.14550.

Dumitrescu, E.-I., & Hurlin, C. (2012). Testing for granger non-causality in heterogeneous panels. Economic modelling, 29(4), 1450–1460.

Fahimi, A., Saint Akadiri, S., Seraj, M., & Akadiri, A. C. (2018). Testing the role of tourism and human capital development in economic growth. a panel causality study of micro states. Tourism management perspectives, 28, 62–70.

Gholipour, H. F., Tajaddini, R., & Farzanegan, M. R. (2023). Governments’ economic support for households during the covid-19 pandemic and consumer confidence. Empirical economics, 65(3), 1253–1272.

Hakobyan, S., Rawlings, H., & Yao, J. (2022). Equitable access to vaccines: Myth or reality?

Hale, T., Angrist, N., Goldszmidt, R., Kira, B., Petherick, A., Phillips, T., … others (2021). A global panel database of pandemic policies (oxford covid-19 government response tracker). Nature human behaviour, 5(4), 529–538.

Hale, T., Angrist, N., Kira, B., Petherick, A., Phillips, T., & Webster, S. (2020). Variation in government responses to covid-19.

Herby, J., Jonung, L., & Hanke, S. (2023). A literature review and meta-analysis of the effects of lockdowns on covid-19 mortality-ii. medRxiv, 2023–08.

Herwartz, H., Maxand, S., Raters, F. H., & Walle, Y. M. (2018). Panel unit-root tests for heteroskedastic panels. The Stata Journal, 18(1), 184–196.

Ji, C., Jiang, J., & Zhang, Y. (2024). Political trust and government performance in the time of covid-19. World Development, 176, 106499.

Kapetanios, G., Pesaran, M. H., & Yamagata, T. (2011). Panels with non-stationary multifactor error structures. Journal of econometrics, 160(2), 326–348.

Koenker, R., & Bassett Jr, G. (1978). Regression quantiles. Econometrica: journal of the Econometric Society, 33–50.

Malki, Z., Atlam, E.-S., Hassanien, A. E., Dagnew, G., Elhosseini, M. A., & Gad, I. (2020). Association between weather data and covid-19 pandemic predicting mortality rate: Machine learning approaches. Chaos, Solitons & Fractals, 138, 110137.

Melly, B., & Pons, M. (n.d.). Minimum distance estimation of quantile panel data models (Tech. Rep.).

Mukherjee, T., Banerjee, A., Mitra, S., & Mukherjee, T. (2022). COVID-19. In Data Science for COVID-19 (pp. 705–728). Elsevier. Retrieved 2023-01-27, from https://linkinghub.elsevier.com/retrieve/pii/B9780323907699000359 doi: 10.1016/B978-0-323-90769-9.00035-9

Namahoro, J., Wu, Q., & Su, H. (2023). Wind energy, industrial-economic development and co2 emissions nexus: Do droughts matter? Energy, 278, 127869.

Nijman, J. (2012). India’s urban challenge. Eurasian Geography and Economics, 53(1), 7–20.

Pesaran, M. H. (2003). Estimation and inference in large heterogenous panels with cross section dependence. Available at SSRN 385123.

Pesaran, M. H. (2007). A simple panel unit root test in the presence of cross-section dependence. Journal of applied econometrics, 22(2), 265–312.

Pesaran, M. H. (2015). Testing weak cross-sectional dependence in large panels. Econometric reviews, 34(6-10), 1089–1117.

Pesaran, M. H. (2021). General diagnostic tests for cross-sectional dependence in panels. Empirical economics, 60(1), 13–50.

Pesaran, M. H., & Yamagata, T. (2008). Testing slope homogeneity in large panels. Journal of econometrics, 142(1), 50–93.

Peters, L. L., Raymer, D. S., Pal, J. D., & Ambardekar, A. V. (2022). Association of covid-19 vaccination with risk of covid-19 infection, hospitalization, and death in heart transplant recipients. JAMA cardiology, 7 (6), 651–654.

Petherick, A., Goldszmidt, R., Andrade, E. B., Furst, R., Hale, T., Pott, A., & Wood, A. (2021). A worldwide assessment of changes in adherence to covid-19 protective behaviours and hypothesized pandemic fatigue. Nature Human Behaviour, 5(9), 1145–1160.

Ritchie, H., Roser, M., & Rosado, P. (2020). Co and greenhouse gas emissions. Our World in Data. (https://ourworldindata.org/co2-and-greenhouse-gas-emissions)

Ritchie, H., Roser, M., & Rosado, P. (2022). Energy. Our World in Data. (https://ourworldindata.org/energy)

Shereen, M. A., Khan, S., Kazmi, A., Bashir, N., & Siddique, R. (2020). Covid-19 infection: Emergence, transmission, and characteristics of human coronaviruses. Journal of advanced research, 24, 91–98.

Sparke, M., & Levy, O. (2022). Competing responses to global inequalities in access to covid vaccines: vaccine diplomacy and vaccine charity versus vaccine liberty. Clinical infectious diseases, 75(Supplement 1), S86–S92.

Stein, C., Nassereldine, H., Sorensen, R. J., Amlag, J. O., Bisignano, C., Byrne, S., … others (2023). Past sars-cov-2 infection protection against re-infection: a systematic review and meta-analysis. The Lancet, 401(10379), 833–842.

Swamy, P. A. (1970). Efficient inference in a random coefficient regression model. Econometrica: Journal of the Econometric Society, 311–323.

Taylor, D., & Shrimankar, D. (2024). The centre and the states: Evolution of a union. In The territories and states of india 2024 (pp. 3–14). Routledge.

Tenforde, M. W., Self, W. H., Adams, K., Gaglani, M., Ginde, A. A., McNeal, T., … others (2021). Association between mrna vaccination and covid-19 hospitalization and disease severity. Jama, 326(20), 2043–2054.

Thiagarajan, K. (2021). Why is india having a covid-19 surge? British Medical Journal Publishing Group.

Varshney, D., Kumar, A., Mishra, A. K., Rashid, S., & Joshi, P. K. (2021). India’s covid-19 social assistance package and its impact on the agriculture sector. Agricultural Systems, 189, 103049.

Wang, M. L., Behrman, P., Dulin, A., Baskin, M. L., Buscemi, J., Alcaraz, K. I., … Fitzgibbon, M. (2020). Addressing inequities in covid-19 morbidity and mortality: research and policy recommendations. Translational Behavioral Medicine, 10(3), 516–519.

Westerlund, J. (2007). Error correction based panel cointegration tests. Oxf Bull Econ Stat, 69, 709–748.

Westerlund, J., & Edgerton, D. L. (2007). A panel bootstrap cointegration test. Economics letters, 97 (3), 185–190.

Westerlund, J., Hosseinkouchack, M., & Solberger, M. (2016). The local power of the cadf and cips panel unit root tests. Econometric Reviews, 35(5), 845–870.

