## Supplementary material for "Assessing the Impact of Non-Pharmaceutical Interventions on COVID-19: A Combined CCE and Quantile Regression Approach"

Kaibalyapati Mishra

July 15, 2024

As I examine the impact of NPIs on COVID-19 outcomes in India, focusing on infection and deaths, the independent variables representing NPIs include stringency measures ($\psi_s$), containment measures ($\psi_c$), economic support measures ($\psi_e$), and overall government support measures ($\psi_o$), which is an aggregated index of the other three. The dependent variables are the weekly infection rates ($I_t$) and weekly death rates ($D_t$). Government spending on these measures can be represented as:

$$G_t = \psi_s \cdot C_s + \psi_c \cdot C_c + \psi_e \cdot C_e$$

where $C_s, C_c, C_e$ are the respective costs associated with each type of measure. We don't mention $C_o$ here though we use it in empirical estimation because $C_s, C_c, C_e$ combined is $C_o$. Households derive utility from both consumption and health levels. The utility function is specified as:

$$U(C_{in}, C_{out}, H) = \alpha \ln(C_{in}) + \beta \ln(C_{out}) + \gamma H$$

where $C_{in}$ denotes indoor consumption, $C_{out}$ denotes outdoor consumption, and $H$ represents the health level of the household. The health level is influenced by the infection rate ($I_t$) and the death rate ($D_t$), with the formulation $H_t = H_0 - \delta I_t - \lambda D_t$. This setup captures the trade-off households face between maintaining consumption activities and protecting their health during the pandemic.

To account for the lag between the implementation of government policies and their observed effects, the model employs dynamic programming. The value function $V_t(S_t)$ is maximized over the policy actions $\psi$, considering both current utility and the expected future utility. The Bellman equation is given by:

$$V_t(S_t) = \max_{\psi} \left[\alpha \ln(C_{in}) + \beta \ln(C_{out}) + \gamma H + \beta \mathbb{E}[V_{t+1}(S_{t+1}) | S_t, \psi_t]\right]$$

The state transition incorporates the time lag $\tau$, with the transition equation $S_{t+\tau} = f(S_t, \psi_t, \epsilon_t)$, where $\epsilon_t$ is a random error term.

The model defines the state transition probabilities to capture the impact of NPIs on infection and death rates. The probability of transitioning from one state $S_t$ to another $S_{t+1}$ is modeled as $\mathbb{P}(S_{t+1} | S_t, \psi_t) = \mathbb{P}(I_{t+1}, D_{t+1} | I_t, D_t, \psi_t)$. The infection dynamics are expressed as $I_{t+1} = I_t \cdot (1 + g(\psi_s, \psi_c, \epsilon_{I,t}))$, and the mortality dynamics are formulated as $D_{t+1} = D_t + h(\psi_s, \psi_c, \psi_e, \psi_o, \epsilon_{D,t})$.

The optimization problem faced by households is to choose the optimal levels of indoor and outdoor activities under the given NPIs to maximize their utility. This is represented as:

$$\max_{\{C_{in}, C_{out}\}} \left\{\alpha \ln(C_{in}) + \beta \ln(C_{out}) + \gamma \left(H_0 - \delta I_t - \lambda D_t\right)\right\}$$

This formulation integrates the trade-offs between consumption and health, considering the direct impact of NPIs on the infection and death rates, thus providing a comprehensive framework for analyzing the effectiveness of government interventions during the COVID-19 pandemic.

### 1 OxCGRT indices and variables of the study

Table 1: OxCGRT indices and variables of the study

| OxCGRT Indicators | | | Targeted/general | | OxCGRT Indices | | | |
|---|---|---|---|---|---|---|---|---|
| ID | Name | Type | | | GRI | CNI | STI | ESI |
| C1 | School closing | Ordinal | Geographic | C1 | x | x | x | |
| C2 | Workplace closing | Ordinal | Geographic | C2 | x | x | x | |
| C3 | Cancel public events | Ordinal | Geographic | C3 | x | x | x | |
| C4 | Restrictions on gathering size | Ordinal | Geographic | C4 | x | x | x | |
| C5 | Close public transport | Ordinal | Geographic | C5 | x | x | x | |
| C6 | Stay-at-home requirements | Ordinal | Geographic | C6 | x | x | x | |
| C7 | Restrictions on internal movement | Ordinal | Geographic | C7 | x | x | x | |
| C8 | Restrictions on international travel | Ordinal | No | C8 | x | x | x | |
| Economic response | | | | E1 | x | | | x |
| E1 | Income support | Ordinal | Sectoral | E2 | x | | | x |
| E2 | Debt/contract relief for households | Ordinal | No | E3 | | | | |
| E3 | Fiscal measures | Numerical | No | E4 | | | | |
| E4 | Giving international support | Numerical | No | H1 | x | x | x | |
| Health systems | | | | H2 | x | x | | |
| H1 | Public information campaign | Ordinal | Geographic | H3 | x | x | | |
| H2 | Testing policy | Ordinal | No | H4 | | | | |
| H3 | Contact tracing | Ordinal | No | H5 | | | | |
| H4 | Emergency investment in health care | Numerical | No | H6 | x | x | | |
| H5 | Investment in COVID-19 vaccines | Numerical | No | H7 | x | x | | |
| H6 | Facial coverings | Ordinal | Geographic | H8 | x | x | | |
| H7 | Vaccination policy | Ordinal | Funding | | | | | |
| H8 | Protection of elderly people | Ordinal | Geographic | k | 16 | 14 | 9 | 2 |

### 2 Diagnostic Test Results

Table 2: Summary statistics of variable quantiles

| *QUINTILES* | *Mean* | *p50* | *SD* | *Min* | *Max* | *Skewness* | *Kurtosis* | *VARIABLES* | *QUINTILES* | *Mean* | *p50* | *SD* | *Min* | *Max* | *Skewness* | *Kurtosis* | *VARIABLES* |
|---|---|---|---|---|---|---|---|---|---|---|---|---|---|---|---|---|---|
| Q1 | -0.005 | 0.100 | 0.395 | -3.323 | 0.100 | -4.505 | 25.173 | | Q1 | 2.755 | 3.046 | 1.779 | 0.100 | 4.605 | -0.342 | 1.497 | |
| Q2 | 3.370 | 3.578 | 1.520 | 0.121 | 5.597 | -0.399 | 2.082 | | Q2 | 4.469 | 4.477 | 0.103 | 4.113 | 4.605 | -0.481 | 2.635 | |
| Q3 | 7.267 | 7.386 | 0.815 | 5.598 | 8.464 | -0.345 | 1.940 | | Q3 | 4.186 | 4.240 | 0.247 | 3.478 | 4.552 | -0.671 | 2.510 | |
| Q4 | 9.123 | 9.132 | 0.358 | 8.470 | 9.715 | -0.095 | 1.844 | | Q4 | 4.037 | 4.082 | 0.293 | 3.290 | 4.577 | -0.307 | 2.062 | |
| Q5 | 10.142 | 10.150 | 0.231 | 9.715 | 10.514 | -0.132 | 1.824 | LnDEATHS | Q5 | 3.964 | 4.050 | 0.394 | 2.631 | 4.577 | -0.999 | 3.865 | LnCONTAINMENT |
| Q6 | 10.814 | 10.820 | 0.163 | 10.515 | 11.078 | -0.118 | 1.807 | | Q6 | 3.748 | 4.050 | 0.654 | 2.408 | 4.577 | -0.786 | 2.040 | |
| Q7 | 11.310 | 11.307 | 0.133 | 11.079 | 11.545 | 0.016 | 1.805 | | Q7 | 3.365 | 3.520 | 0.726 | 2.408 | 4.477 | 0.041 | 1.261 | |
| Q8 | 11.795 | 11.784 | 0.151 | 11.545 | 12.069 | 0.107 | 1.808 | | Q8 | 3.380 | 3.561 | 0.698 | 2.408 | 4.577 | -0.018 | 1.378 | |
| Q9 | 12.349 | 12.345 | 0.165 | 12.070 | 12.655 | 0.104 | 1.861 | | Q9 | 3.069 | 2.631 | 0.641 | 2.408 | 4.561 | 0.816 | 2.026 | |
| Q10 | 13.192 | 13.101 | 0.410 | 12.655 | 14.280 | 0.601 | 2.371 | | Q10 | 2.901 | 2.631 | 0.579 | 2.408 | 4.293 | 1.417 | 3.321 | |
| Q1 | 1.632 | 0.100 | 2.824 | -2.388 | 11.693 | 1.638 | 4.831 | | Q1 | 2.547 | 2.604 | 1.805 | -0.265 | 4.531 | -0.193 | 1.337 | |
| Q2 | 7.994 | 8.149 | 2.034 | 2.208 | 12.698 | -0.178 | 2.582 | | Q2 | 4.398 | 4.404 | 0.067 | 4.145 | 4.531 | -0.454 | 2.848 | |
| Q3 | 11.987 | 12.116 | 1.167 | 8.690 | 14.299 | -0.406 | 2.593 | | Q3 | 4.200 | 4.235 | 0.175 | 3.758 | 4.485 | -0.567 | 2.266 | |
| Q4 | 13.716 | 13.714 | 0.675 | 11.740 | 15.310 | -0.096 | 2.876 | | Q4 | 4.134 | 4.135 | 0.200 | 3.628 | 4.563 | -0.102 | 2.224 | |
| Q5 | 14.611 | 14.572 | 0.541 | 13.187 | 16.174 | 0.315 | 3.185 | LnINFECTIONS | Q5 | 4.123 | 4.142 | 0.228 | 3.470 | 4.563 | -0.533 | 2.945 | LnSTRINGENCY |
| Q6 | 15.311 | 15.237 | 0.483 | 13.999 | 16.692 | 0.433 | 3.223 | | Q6 | 4.024 | 4.154 | 0.340 | 3.288 | 4.537 | -0.628 | 1.942 | |
| Q7 | 15.817 | 15.736 | 0.447 | 14.681 | 17.174 | 0.549 | 3.315 | | Q7 | 3.835 | 3.863 | 0.367 | 3.288 | 4.486 | 0.112 | 1.469 | |
| Q8 | 16.222 | 16.164 | 0.449 | 15.133 | 17.765 | 0.836 | 4.137 | | Q8 | 3.853 | 3.876 | 0.354 | 3.145 | 4.548 | -0.004 | 1.715 | |
| Q9 | 16.668 | 16.619 | 0.450 | 15.658 | 18.313 | 1.316 | 5.974 | | Q9 | 3.693 | 3.524 | 0.327 | 3.145 | 4.537 | 0.802 | 2.396 | |
| Q10 | 17.392 | 17.332 | 0.492 | 16.323 | 18.469 | 0.209 | 2.293 | | Q10 | 3.600 | 3.470 | 0.295 | 3.145 | 4.309 | 1.222 | 3.288 | |
| Q1 | 2.499 | 2.543 | 1.819 | -0.403 | 4.507 | -0.156 | 1.307 | | Q1 | 1.941 | 0.100 | 2.060 | 0.100 | 4.317 | 0.240 | 1.079 | |
| Q2 | 4.386 | 4.391 | 0.060 | 4.168 | 4.507 | -0.451 | 2.833 | | Q2 | 4.295 | 4.317 | 0.060 | 4.135 | 4.317 | -2.278 | 6.227 | |
| Q3 | 4.212 | 4.238 | 0.150 | 3.847 | 4.466 | -0.525 | 2.186 | | Q3 | 4.274 | 4.317 | 0.162 | 3.219 | 4.317 | -5.086 | 30.957 | |
| Q4 | 4.141 | 4.152 | 0.166 | 3.704 | 4.501 | -0.230 | 2.421 | | Q4 | 4.101 | 4.317 | 0.364 | 2.526 | 4.317 | -1.791 | 5.771 | |
| Q5 | 4.115 | 4.136 | 0.202 | 3.491 | 4.501 | -0.736 | 3.510 | LnOVERALL | Q5 | 3.982 | 3.912 | 0.323 | 3.219 | 4.317 | -0.855 | 3.257 | LnECONOMIC |
| Q6 | 3.991 | 4.127 | 0.341 | 3.219 | 4.466 | -0.764 | 2.121 | | Q6 | 3.606 | 3.912 | 0.762 | 0.100 | 4.317 | -3.059 | 14.285 | |
| Q7 | 3.788 | 3.831 | 0.391 | 3.154 | 4.430 | -0.069 | 1.485 | | Q7 | 2.813 | 3.912 | 1.637 | 0.100 | 4.317 | -0.971 | 2.116 | |
| Q8 | 3.802 | 3.843 | 0.372 | 3.011 | 4.510 | -0.115 | 1.743 | | Q8 | 2.917 | 3.219 | 1.461 | 0.100 | 4.317 | -1.302 | 2.941 | |
| Q9 | 3.621 | 3.459 | 0.349 | 3.011 | 4.513 | 0.761 | 2.396 | | Q9 | 2.210 | 3.219 | 1.647 | 0.100 | 4.605 | -0.432 | 1.343 | |
| Q10 | 3.512 | 3.357 | 0.332 | 3.011 | 4.279 | 1.155 | 3.108 | | Q10 | 1.601 | 0.100 | 1.700 | 0.100 | 4.605 | 0.333 | 1.287 | |

#### 2.1 Results

Table 3: Results from quantile regression analysis with Deaths due to COVID-19 as dependent variable (Q1 to Q3)

| ***d.LnINFECTIONS*** | **Overall** | | **Stringency** | | **Containment** | | **Economic** | |
|---|---|---|---|---|---|---|---|---|
| | **(1)** | | **(2)** | | **(3)** | | **(4)** | |
| | | | | | **Q1** | | | |
| *LnINFECTIONS* | 0.973** | (0.0102) | 0.867* | (0.00672) | 0.986* | (0.0111) | 1.064** | (0.0211) |
| *LnOVERALL* | -1.428*** | (0.0465) | | | | | | |
| *LnSTRINGENCY* | | | -1.067*** | (0.0358) | | | | |
| *LnCONTAINMENT* | | | | | -1.471*** | (0.0494) | | |
| *LnECONOMIC* | | | | | | | -0.129*** | (0.0182) |
| *Constant* | 0.145* | (0.0493) | -0.206* | (0.0143) | 0.148* | (0.00853) | -6.240** | (0.364) |
| | | | | | ***Q2*** | | | |
| *LnINFECTIONS* | 0.922** | (0.00466) | 0.843* | (0.00364) | 0.928* | (0.00558) | 0.919** | (0.0101) |
| *LnOVERALL* | -1.055*** | (0.0204) | | | | | | |
| *LnSTRINGENCY* | | | -0.845*** | (0.0183) | | | | |
| *LnCONTAINMENT* | | | | | -1.069*** | (0.0239) | | |
| *LnECONOMIC* | | | | | | | -0.244*** | (0.0114) |
| *Constant* | 0.113** | (0.0330) | 0.100* | (0.0135) | 0.114* | (0.0317) | -3.176** | (0.172) |
| | | | | | ***Q3*** | | | |
| *LnINFECTIONS* | 0.898** | (0.00492) | 0.825* | (0.00366) | 0.905* | (0.00560) | 0.885** | (0.0108) |
| *LnOVERALL* | -0.882*** | (0.0214) | | | | | | |
| *LnSTRINGENCY* | | | -0.670*** | (0.0167) | | | | |
| *LnCONTAINMENT* | | | | | -0.900*** | (0.0241) | | |
| *LnECONOMIC* | | | | | | | -0.183*** | (0.0128) |
| *Constant* | 0.0984** | (0.0279) | 0.0845 | (0.0339) | 0.0995** | (0.0283) | -2.520** | (0.188) |
| *N* | 5148 | | 5148 | | 5148 | | 5148 | |

*Standard errors in parentheses — =* $^{*}$ *p<0.05,* $^{**}$ *p<0.01,* $^{***}$ *p<0.001*

Table 4: Results from quantile regression analysis with Deaths due to COVID-19 as dependent variable (Q4 to Q6)

| ***d.LnINFECTIONS*** | **Overall** | | **Stringency** | | **Containment** | | **Economic** | |
|---|---|---|---|---|---|---|---|---|
| | **(1)** | | **(2)** | | **(3)** | | **(4)** | |
| | | | | ***Q4*** | | | | |
| *LnINFECTIONS* | 0.875** | (0.00387) | 0.817* | (0.00298) | 0.881* | (0.00454) | 0.844** | (0.00948) |
| *LnOVERALL* | -0.726*** | (0.0159) | | | | | | |
| *LnSTRINGENCY* | | | -0.567*** | (0.0123) | | | | |
| *LnCONTAINMENT* | | | | | -0.747*** | (0.0188) | | |
| *LnECONOMIC* | | | | | | | -0.195*** | (0.0122) |
| *Constant* | 0.0852** | (0.0161) | 0.0750 | (0.0319) | 0.0866** | (0.0251) | -1.673** | (0.170) |
| | | | | ***Q5*** | | | | |
| *LnINFECTIONS* | 0.866** | (0.00326) | 0.814* | (0.00322) | 0.871* | (0.00360) | 0.789** | (0.0118) |
| *LnOVERALL* | -0.649*** | (0.0129) | | | | | | |
| *LnSTRINGENCY* | | | -0.489*** | (0.0128) | | | | |
| *LnCONTAINMENT* | | | | | -0.657*** | (0.0144) | | |
| *LnECONOMIC* | | | | | | | -0.219*** | (0.0134) |
| *Constant* | 0.0783** | (0.0235) | 0.0675 | (0.0297) | 0.0786** | (0.0212) | -0.600* | (0.212) |
| | | | | ***Q6*** | | | | |
| *LnINFECTIONS* | 0.851** | (0.00387) | 0.805* | (0.00340) | 0.858* | (0.00421) | 0.760** | (0.00182) |
| *LnOVERALL* | -0.577*** | (0.0155) | | | | | | |
| *LnSTRINGENCY* | | | -0.397*** | (0.0124) | | | | |
| *LnCONTAINMENT* | | | | | -0.568*** | (0.0162) | | |
| *LnECONOMIC* | | | | | | | -0.229*** | (0.00719) |
| *Constant* | 0.182** | (0.0229) | 0.0592 | (0.0337) | 0.0711 | (0.0245) | 0.0469** | (0.0141) |
| *N* | 5148 | | 5148 | | 5148 | | 5148 | |

*Standard errors in parentheses — =* $^{*}$ $p<0.05$, $^{**}$ $p<0.01$, $^{***}$ $p<0.001$

Table 5: Results from quantile regression analysis with Deaths due to COVID-19 as dependent variable (Q7 to Q10)

| ***d.LnINFECTIONS*** | **Overall** | | **Stringency** | | **Containment** | | **Economic** | |
|---|---|---|---|---|---|---|---|---|
| | **(1)** | | **(2)** | | **(3)** | | **(4)** | |
| | | | | | ***Q7*** | | | |
| *LnINFECTIONS* | 0.836** | (0.00400) | 0.797* | (0.00353) | 0.837* | (0.00465) | 0.766** | (0.00211) |
| *LnOVERALL* | -0.498*** | (0.0159) | | | | | | |
| *LnSTRINGENCY* | | | -0.326*** | (0.0122) | | | | |
| *LnCONTAINMENT* | | | | | -0.514*** | (0.0184) | | |
| *LnECONOMIC* | | | | | | | -0.206*** | (0.00769) |
| *Constant* | 0.238** | (0.0239) | 0.0774 | (0.0285) | 0.315* | (0.0289) | 0.0440 | (0.0186) |
| | | | | | ***Q8*** | | | |
| *LnINFECTIONS* | 0.814** | (0.00437) | 0.774* | (0.00344) | 0.818* | (0.00476) | 0.767** | (0.00187) |
| *LnOVERALL* | -0.456*** | (0.0192) | | | | | | |
| *LnSTRINGENCY* | | | -0.273*** | (0.0137) | | | | |
| *LnCONTAINMENT* | | | | | -0.426*** | (0.0182) | | |
| *LnECONOMIC* | | | | | | | -0.165*** | (0.00759) |
| *Constant* | 0.536** | (0.0692) | 0.370* | (0.0301) | 0.373* | (0.0339) | 0.0398** | (0.0107) |
| | | | | | ***Q9*** | | | |
| *LnINFECTIONS* | 0.788** | (0.00462) | 0.766* | (0.00302) | 0.790* | (0.00515) | 0.772** | (0.00211) |
| *LnOVERALL* | -0.316*** | (0.0246) | | | | | | |
| *LnSTRINGENCY* | | | -0.217*** | (0.0144) | | | | |
| *LnCONTAINMENT* | | | | | -0.324*** | (0.0281) | | |
| *LnECONOMIC* | | | | | | | -0.140*** | (0.00850) |
| *Constant* | 0.563** | (0.0456) | 0.484* | (0.0371) | 0.594* | (0.0472) | 0.0369** | (0.0103) |
| | | | | | ***Q10*** | | | |
| *LnINFECTIONS* | 0.756** | (0.00430) | 0.752* | (0.00293) | 0.757* | (0.00444) | 0.759** | (0.00158) |
| *LnOVERALL* | -0.280*** | (0.0373) | | | | | | |
| *LnSTRINGENCY* | | | -0.176*** | (0.0153) | | | | |
| *LnCONTAINMENT* | | | | | -0.272*** | (0.0426) | | |
| *LnECONOMIC* | | | | | | | -0.0663*** | (0.00543) |
| *Constant* | 1.252** | (0.183) | 0.831* | (0.0765) | 1.220* | (0.207) | 0.310** | (0.0200) |
| *N* | 5148 | | 5148 | | 5148 | | 5148 | |

*Standard errors in parentheses — =* $p<0.05$, ** $p<0.01$, *** $p<0.001$*

Table 6: Results from quantile regression analysis with Infections of COVID-19 as dependent variable (Q1 to Q4)

| ***d.LnINFECTIONS*** | **Overall** | | **Stringency** | | **Containment** | | **Economic** | |
|---|---|---|---|---|---|---|---|---|
| | **(5)** | | **(6)** | | **(7)** | | **(8)** | |
| **Q1** | | | | | | | | |
| *LnOVERALL* | 1.447*** | (0.0881) | | | | | | |
| *LnSTRINGENCY* | | | 1.119*** | (0.0918) | | | | |
| *LnCONTAINMENT* | | | | | 1.434*** | (0.0905) | | |
| *LnECONOMIC* | | | | | | | 0.967*** | (0.0687) |
| *Constant* | -2.552** | (0.200) | -2.272* | (0.232) | -2.558** | (0.206) | 0.00332 | (0.120) |
| ***Q2*** | | | | | | | | |
| *LnOVERALL* | 2.422*** | (0.0544) | | | | | | |
| *LnSTRINGENCY* | | | 1.997*** | (0.0734) | | | | |
| *LnCONTAINMENT* | | | | | 2.501*** | (0.0616) | | |
| *LnECONOMIC* | | | | | | | 2.183*** | (0.0568) |
| *Constant* | -0.977** | (0.142) | -0.0997 | (0.259) | -1.356** | (0.165) | -0.118 | (0.134) |
| ***Q3*** | | | | | | | | |
| *LnOVERALL* | 3.074*** | (0.0470) | | | | | | |
| *LnSTRINGENCY* | | | -2.031*** | (0.179) | | | | |
| *LnCONTAINMENT* | | | | | 3.087*** | (0.0492) | | |
| *LnECONOMIC* | | | | | | | -0.801*** | (0.0435) |
| *Constant* | -0.207 | (0.173) | 20.40** | (0.638) | -0.209 | (0.185) | 15.53** | (0.145) |
| ***Q4*** | | | | | | | | |
| *LnOVERALL* | 3.369*** | (0.0415) | | | | | | |
| *LnSTRINGENCY* | | | -1.830*** | (0.100) | | | | |
| *LnCONTAINMENT* | | | | | 3.364*** | (0.0442) | | |
| *LnECONOMIC* | | | | | | | -0.680*** | (0.0232) |
| *Constant* | -0.237 | (0.168) | 20.58** | (0.323) | -0.236 | (0.178) | 16.22** | (0.0690) |
| *N* | 5148 | | 5148 | | 5148 | | 5148 | |

*Standard errors in parentheses — =* $p<0.05$, ** $p<0.01$, *** $p<0.001$*

Table 7: Results from quantile regression analysis with Infections of COVID-19 as dependent variable (Q5 to Q8)

| ***d.LnINFECTIONS*** | **Overall** | | **Stringency** | | **Containment** | | **Economic** | |
|---|---|---|---|---|---|---|---|---|
| | **(5)** | | **(6)** | | **(7)** | | **(8)** | |
| | | | | ***Q5*** | | | | |
| *LnOVERALL* | 1.730 | (1.537) | | | | | | |
| *LnSTRINGENCY* | | | -1.508*** | (0.0763) | | | | |
| *LnCONTAINMENT* | | | | | 2.999*** | (0.354) | | |
| *LnECONOMIC* | | | | | | | -0.589*** | (0.0197) |
| *_cons* | 7.432 | (6.372) | 20.06** | (0.248) | 2.138 | (1.483) | 16.57** | (0.0596) |
| | | | | ***Q6*** | | | | |
| *LnOVERALL* | -0.882*** | (0.186) | | | | | | |
| *LnSTRINGENCY* | | | -1.279*** | (0.0596) | | | | |
| *LnCONTAINMENT* | | | | | -0.577* | (0.232) | | |
| *LnECONOMIC* | | | | | | | -0.534*** | (0.0185) |
| *Constant* | 18.77** | (0.728) | 19.73* | (0.196) | 17.59* | (0.923) | 16.89** | (0.0569) |
| | | | | ***Q7*** | | | | |
| *LnOVERALL* | -1.158*** | (0.130) | | | | | | |
| *LnSTRINGENCY* | | | -1.065*** | (0.0471) | | | | |
| *LnCONTAINMENT* | | | | | -1.005*** | (0.154) | | |
| *LnECONOMIC* | | | | | | | -0.465*** | (0.0171) |
| *Constant* | 20.24** | (0.501) | 19.45* | (0.159) | 19.72* | (0.603) | 17.09** | (0.0533) |
| | | | | ***Q8*** | | | | |
| *LnOVERALL* | -1.292*** | (0.100) | | | | | | |
| *LnSTRINGENCY* | | | -0.988*** | (0.0406) | | | | |
| *LnCONTAINMENT* | | | | | -1.144*** | (0.114) | | |
| *LnECONOMIC* | | | | | | | -0.412*** | (0.0166) |
| *Constant* | 21.10** | (0.385) | 19.55* | (0.138) | 20.60* | (0.445) | 17.35** | (0.0511) |
| *N* | 5148 | | 5148 | | 5148 | | 5148 | |

*Standard errors in parentheses — =* $^{*}$ $p<0.05$, $^{**}$ $p<0.01$, $^{***}$ $p<0.001$

Table 8: Results from quantile regression analysis with Infections of COVID-19 as dependent variable (Q9 to Q10)

| ***d.LnINFECTIONS*** | **Overall** (5) | | **Stringency** (6) | | **Containment** (7) | | **Economic** (8) | |
|---|---|---|---|---|---|---|---|---|
| | ***Q9*** | | | | | | | |
| *LnOVERALL* | -1.207*** | (0.0793) | | | | | | |
| *LnSTRINGENCY* | | | -0.970*** | (0.0365) | | | | |
| *LnCONTAINMENT* | | | | | -1.160*** | (0.0891) | | |
| *LnECONOMIC* | | | | | | | -0.434*** | (0.0176) |
| *Constant* | 21.09** | (0.304) | 19.85* | (0.126) | 21.03* | (0.344) | 17.88** | (0.0572) |
| | ***Q10*** | | | | | | | |
| *LnOVERALL* | -1.303*** | (0.0766) | | | | | | |
| *LnSTRINGENCY* | | | -0.916*** | (0.0387) | | | | |
| *LnCONTAINMENT* | | | | | -1.334*** | (0.0798) | | |
| *LnECONOMIC* | | | | | | | -0.349*** | (0.0146) |
| *Constant* | 22.27** | (0.293) | 20.38* | (0.135) | 22.47* | (0.308) | 18.22** | (0.0457) |
| *N* | 5148 | | 5148 | | 5148 | | 5148 | |

*Standard errors in parentheses — =* $^{*}$ $p<0.05$, $^{**}$ $p<0.01$, $^{***}$ $p<0.001$
